# Statistical methods for chemical mixtures: a roadmap for practitioners

**DOI:** 10.1101/2024.03.03.24303677

**Authors:** Wei Hao, Amber L. Cathey, Max M. Aung, Jonathan Boss, John D. Meeker, Bhramar Mukherjee

## Abstract

Quantitative characterization of the health impacts associated with exposure to chemical mixtures has received considerable attention in current environmental and epidemiological studies. With many existing statistical methods and emerging approaches, it is important for practitioners to understand when each method is best suited for their inferential goals. In this study, we conduct a review and comparison of 11 analytical methods available for use in mixtures research, through extensive simulation studies for continuous and binary outcomes. These methods fall in three different classes: identifying important components of a mixture, identifying interactions and creating a summary score for risk stratification and prediction. We carry out an illustrative data analysis in the PROTECT birth cohort from Puerto Rico. Most importantly we develop an integrated package “CompMix” that provides a platform for mixtures analysis where the practitioner can implement a pipeline for several types of mixtures analysis.

Our simulation results suggest that the choice of methods depends on the goal of analysis and there is no clear winner across the board. For selection of important toxicants in the mixture and for identifying interactions, Elastic net by Zou et al. (Enet), Lasso for Hierarchical Interactions by Bien et al (HierNet), Selection of nonlinear interactions by a forward stepwise algorithm by Narisetty et al. (SNIF) have the most stable performance across simulation settings. Additionally, the predictive performance of the Super Learner ensembling method by Van de Laan et al. and HierNet are found to be superior to the rest of the methods. For overall summary or a cumulative measure, we find that using the Super Learner to combine multiple Environmental Risk Scores can lead to improved risk stratification properties. We have developed an R package “CompMix: A comprehensive toolkit for environmental mixtures analysis”, allowing users to implement a variety of tasks under different settings and compare the findings.

In summary, our study offers guidelines for selecting appropriate statistical methods for addressing specific scientific questions related to mixtures research. We identify critical gaps where new and better methods are needed.

## Introduction

In recent years, many environmental health studies have explored chemical mixtures using a variety of statistical methods aimed at characterizing the mixture and assessing the mixture’s effects on health outcomes. For example, these chemical mixtures or multipollutant may include phthalates, phenols, polycyclic aromatic hydrocarbons (PAHs), per- and polyfluoroalkyl substances (PFAS), metals and more. Traditional studies of health impacts of environmental exposure have focused on examining individual agents one at a time, primarily due to the limitations in statistical methods and prohibitive sample sizes.

However, in reality, humans are exposed to a wide range of chemicals they encounter in their environments via various pathways simultaneously, which poses significant statistical challenges when studying the joint health effect of the mixture. For example, the chemicals may exhibit complex dependence; the response-dose associations are often highly nonlinear and nonadditive; the number of the multipollutant and their potential interactions could be high, with their effect sizes potentially small and challenging to detect compared to the larger effect of demographic covariates. These challenges are difficult to address satisfactorily via standard regression models.

The National Institute of Environmental Health Sciences (NIEHS) has identified mixtures analyses as a high-priority area for research in 2013 and 2015 [1, 2], and launched the Powering Research through Innovative Methods for Mixtures in Epidemiology (PRIME) funding program to address methodological challenges in mixtures research in 2017 [3]. Despite the development and availability of numerous mixtures methods, there is continued discussion and debate on which methods are best suited for a given researcher’s hypothesis for a given data set. The central goal of this paper is to provide empirical evidence regarding performance of mixture methods to help guide researchers on selecting the best available methods to address three scientific questions in data analysis: (1) identifying the important toxic components of the mixture as related to the health outcome; (2) identifying the interaction effects from combinations of pollutants on the outcome; and (3) prediction of the health outcome and identifying high-risk mixture strata.

Most importantly, we want to streamline implementation challenges so that practitioners are able to explore a variety of methods in a single platform. To this end, we have developed an R package “CompMix: A comprehensive toolkit for environmental mixtures analysis”. The package offers the flexibility to perform various tasks such as variable selection, interaction detection, form composite summary risk scores and compare certain performance metrics across fitted models. Our vision going forward is to update CompMix with emerging methods as they become available.

Our study is motivated by a large-scale NIH-funded longitudinal birth cohort study taking place in Puerto Rico known as the PROTECT study, which aims to increase diversity and representation of historically neglected communities in biomedical research and investigate how exposures to a range of chemicals in the environment, including phthalates, phenols, PAHs, and metals, negatively impact birth outcomes and women’s health. The recruitment and study protocols for PROTECT have been previously described [4, 5]. Puerto Rico has 18 Superfund sites and suffers extensively from environmental contamination. At the same time, the population in Puerto Rico has higher rates of preterm birth and low birth weight with 11.6% of live births being preterm and 10.2% being low birthweight, compared to 10.1% and 8.2%, respectively, in the general United States population in 2020 [6]. Adverse birth outcomes, such as preterm birth (less than 37 weeks gestation), and low birth weight (birth weight less than 2500 grams), are global health concerns linked to increased risks of developing conditions such as diabetes and cardiovascular disease in adulthood [7, 8]. Previous studies utilizing the PROTECT cohort have observed links between individual environmental chemical exposures during pregnancy and a greater risk of preterm birth [9]. However, due to unique statistical barriers, much remains unknown about the impact of exposure to environmental toxicant mixtures during pregnancy and these adverse birth outcomes. Figure 1 shows a correlation heatmap of mean log-transformed concentrations across three prenatal visits for 39 chemical exposures from urine samples in the PROTECT study.

**Figure 1:**
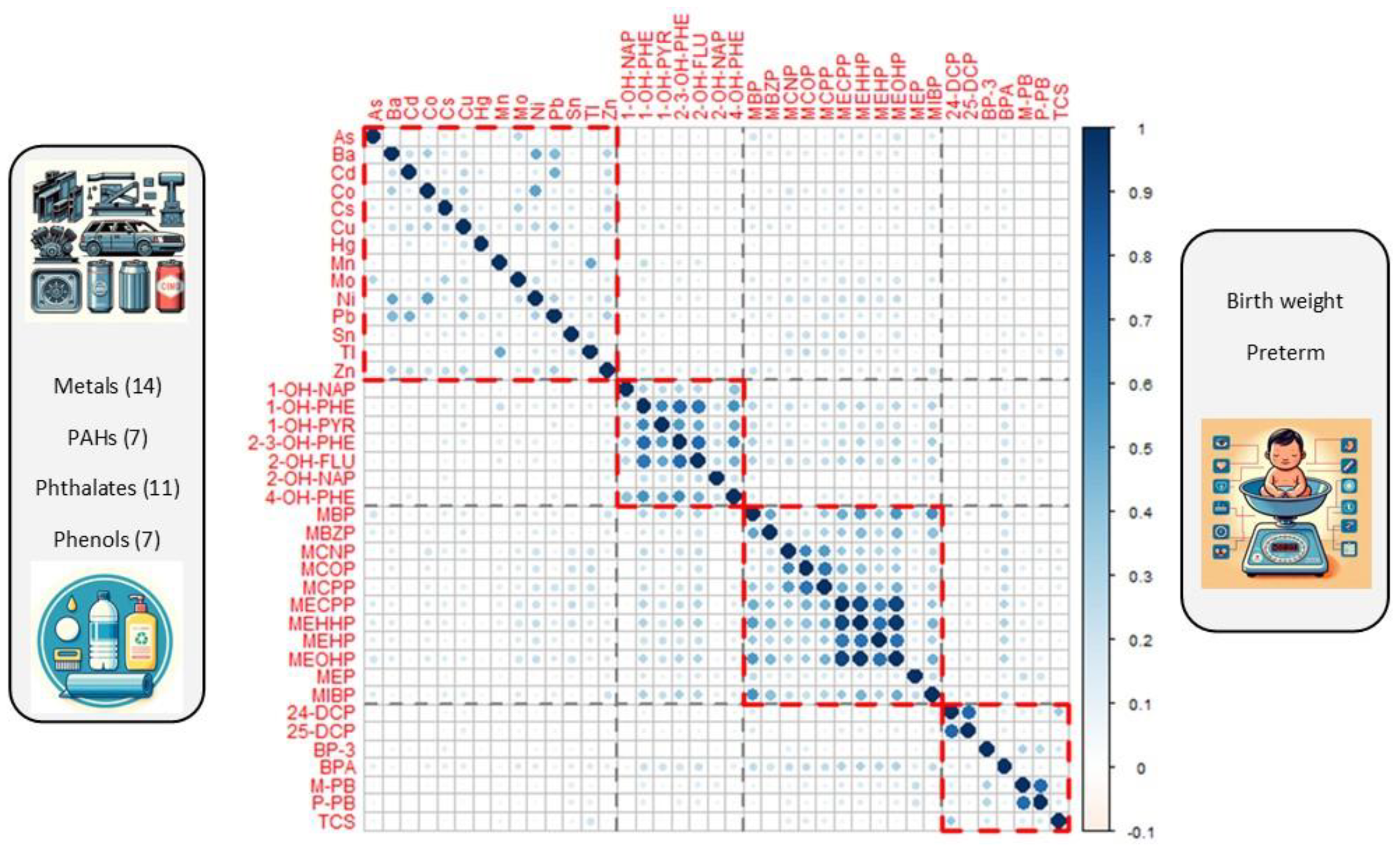
Correlation heatmap of mean concentrations across three prenatal visits for 39 chemicals from urine samples in the PROTECT study, where the concentrations were adjusted for specific gravity and taken logarithm with base 10. The chemicals are ordered by four families: metals, PAHs, phthalates, and phenols.

### The landscape of Statistical Methods

Popular approaches for identifying mixture components are high-dimensional penalized regressions such as Lasso [10], Elastic Net (Enet [11]) and Group Lasso [12]. Other more flexible approaches with nonparametric natures include machine learning methods such as random forest (RF [13]), neural networks and support vector machine. One important goal in mixtures research is to identify interactions among exposures, which motivated the development of hierarchical integrative group least absolute shrinkage and selection operator (Higlasso [14]), selection of nonlinear interactions by a forward stepwise algorithm (SNIF [15]), factor analysis for interactions (FIN [16]), and Bayesian kernel machine regression (BKMR) [17, 18]. Lasso for Hierarchical Interactions (HierNet [19]) is a general method for interaction detection and has also been utilized for the mixtures analysis.

One special area of machine learning is ensemble learning, and its representative work is Super Learner [20] targeted towards optimal prediction, but can also be used for variable identification through creation of an importance score. Moreover, methods have also been developed to characterize the summary measures of environmental mixtures, including weighted quantile sum regression (WQS) [21] and quantile g-computation (Q-gcomp) [22]. In particular, Environmental Risk Score (ERS), a general method that utilizes a diverse range of predictive models to construct a one-dimensional risk score [23, 24], has also attracted attention and has been broadly applied to quantify the health effects due to the pollutant mixtures.

Several publications have provided an overview of many statistical approaches available for studying the health effects of chemical mixtures. Davalos et al. [25] reviewed approaches used in examining air pollution exposures and classified these approaches into five classes. Gibson et al. [26] provided an extensive overview of the methods and illustrated their usage with the National Health and Nutrition Examination Survey (NHANES) dataset. Park et al. [24] focused on the machine learning approaches to construct the ERS with an NHANES data analysis as an illustration. These publications have focused on real data analysis, meaning that one would never know the true contributing toxicants and associations given the data, and making it difficult to evaluate the selection accuracy of the pollutants among methods. In contrast, simulation studies are a powerful tool for comprehensively comparing various methods under a broad range of data generating mechanisms. Some sporadic works on simulation studies for mixtures analysis [27, 28] have emerged, but there still lacks a systematic evaluation of the popular methods used in mixture analyses under diverse data scenarios with varied sample sizes, changing number of pollutants for continuous and binary outcomes. To address this gap, our goal is to utilize simulation studies to perform a head-to-head comparison among mixtures methods under different data settings and provide guidance for practitioners.

The present study proposes an analytical framework that utilizes variable selection/prioritization/ranking techniques including Lasso, Enet, Group Lasso, BKMR, RF, Higlasso, HierNet and SNIF to identify the pollutants and interactions that are associated with the health outcome. We characterize the health effects from exposure to the mixtures as ERS, and adapt the ensemble learning approach of Super Learner [20] to combine the ERSs derived from various methods for improved prediction. We compare the different ERSs with summary measures derived from WQS and Q-gcomp.

The evaluation metrics that we consider in our simulation study include: measures of variable selection accuracy, prediction accuracy under different outcome types, ability to stratify high-risk individuals and the computational cost. By varying the sample size, the number of pollutants, and the signal to noise ratio, we strive to provide a comprehensive evaluation of the representative methods and gain insight into their advantages and limitations in the context of mixtures analysis.

## General framework

We select 11 representative statistical approaches and categorize them into three groups based on their ability to tackle the three main objectives of a typical mixture analysis plan: (1) identifying the important toxic components of the mixture; (2) identifying the interaction effects of combination of pollutants, and (3) evaluating the predictive performance of the summary measures and risk stratification. Importantly, all of these goals are specific to an underlying health outcome. The grouping of methods is presented in Figure 2, and further details of each method can be found in the Methods section.

**Figure 2:**
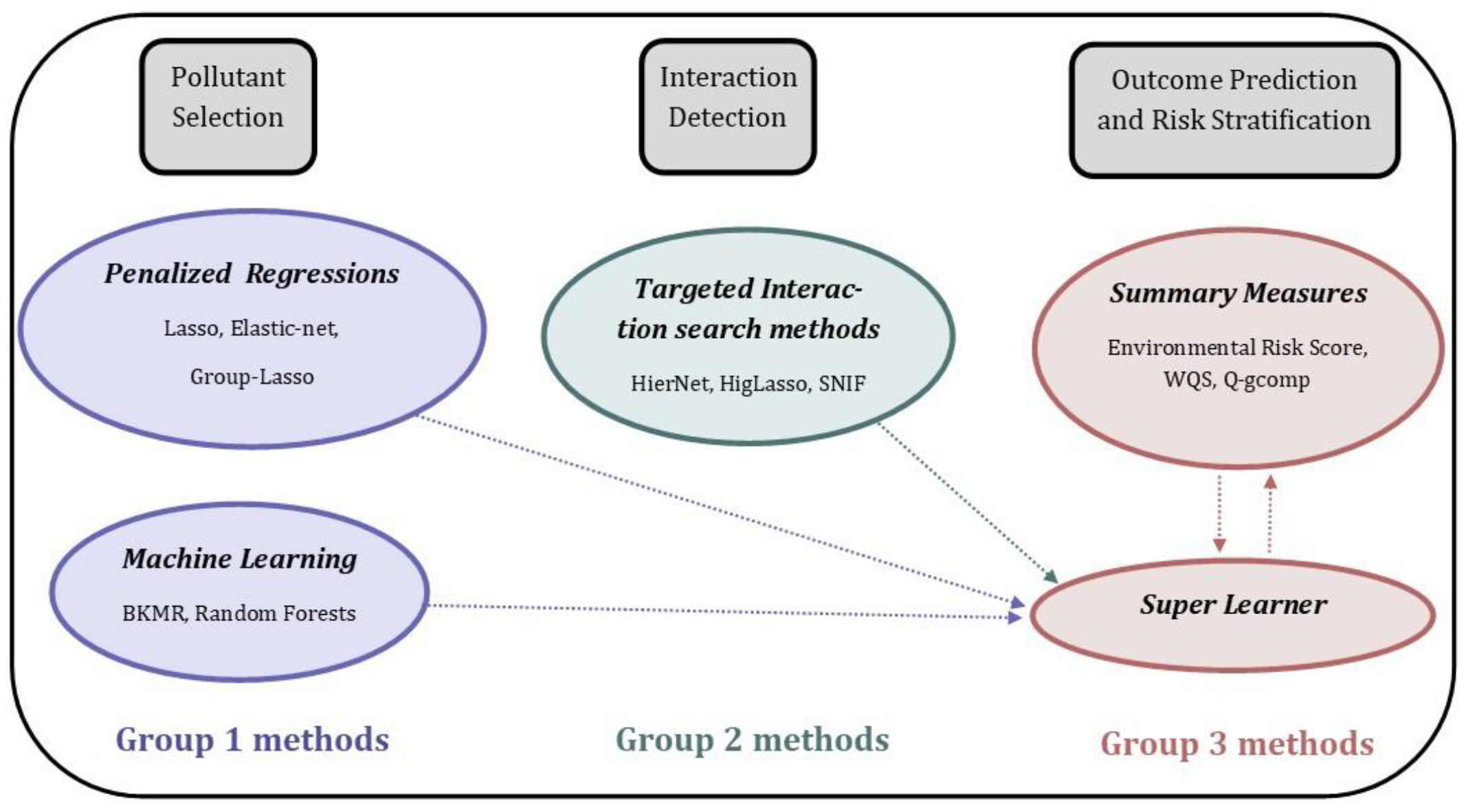
Methods for mixtures analysis categorized in three groups.

For objective 1, which involves pollutant selection, we consider methods that perform variable selection or provide importance scores corresponding to the variables that can be used for ranking and can be thresholded for variable selection. These methods include penalized regressions such as Lasso, Elastic net (Enet), and Group Lasso (G-Lasso). We also consider two machine learning methods that provide rankings of the pollutant importance: BKMR and Random forests (RF).

For objective 2, which involves interaction detection, we will consider three methods that were specifically developed for interaction selection. These methods include Hierarchical Integrative G-Lasso (HigLasso), Lasso for Hierarchical Interactions (HierNet), and Selection of nonlinear interactions by a forward stepwise algorithm (SNIF). HigLasso and SNIF are particularly motived by problems in chemical mixtures analysis. It is worth noting that some of the Group 1 methods, such as Lasso, Enet and G-Lasso can also be used for interaction selection if one specifies the interaction terms in the underlying models.

However, these methods were not initially designed for interaction selection, so they do not account for key assumptions such as heredity principles [14]. For the three methods in Group 2, only the individual pollutants need to be specified; the methods will automatically select the pairwise interactions or even quadratic terms of pollutants (HierNet).

For objective 3, which involves prediction of health outcomes and risk stratification, we will consider four methods, including Environmental Risk Score (ERS), weighted quantile sum regression (WQS), quantile g-computation (Q-gcomp), and Super Learner (SL). ERS utilizes predictive models to create a summary risk score, while WQS and Q-gcomp also construct a summary measure of body burden index from a weighted average of exposures. These three summary measures are used to characterize the joint cumulative health impacts resulting from exposure to mixtures of pollutants, and all serve as dimension reduction measures in the mixture analysis. SL uses cross-validation to create a weighted combination of different learners to improve prediction. Motivated by the fundamental idea of SL, we have developed our own version of SL for ensembling various ERSs. We refer to this method as SL-ERS.

For fair evaluation, the simulated data are split evenly into training and testing sets. In the training data, we compare the performance of pollutant selection and interaction detection for methods in Groups 1 and 2. The evaluation metrics include sensitivity, specificity, false discovery rate and false positive rate. In the testing data, we evaluate the prediction and risk stratification properties of summary measures for methods in Group 3, which include ERSs constructed from several methods that demonstrate good performance for pollutant selection and interaction detection, SL-ERS, WQS and Q-gcomp. The evaluation metrics for continuous outcomes include correlation coefficient (Corr) between ERS (or WQS/Q-gcomp summary scores) and health outcomes and sum of squared error (SSE) corresponding to predicting the health outcome by these risk scores. For creating a binary outcome, we dichotomize the continuous outcome at 90th percentile, and compute the area under the receiver operating characteristic curve (AUC) as a measure of discrimination. Lastly, we stratify each summary risk score measures by the 25 and 75 percentiles (Q_1_ and Q_3_) to create the low- or high-risk groups, fit a logistic regression and report the odds ratio (OR) of having an extreme outcome between the group with the lowest quartile of the summary measure and the group with the highest quartile of the summary measure. The odds ratio serves as the metric for assessing the risk discrimination property of the summary scores. The details of all the evaluation metrics can be found in the Methods section.

## Results

### Simulation Results

We conduct simulation studies to compare the performance of different methods. We consider 20 pollutants representing three families of environmental chemicals, namely phthalates, PAHs, and metals. These pollutants are divided into three groups, with seven, six and seven pollutants in each group, respectively. There are five true features distributed as two, two and one pollutant in each group, respectively. The correlation matrix of pollutant exposures is specified according to the grouping structure, where within-group correlations and between-group correlations are set to 0.6 and 0.1. Table 8 presents the complete list of simulation settings. The mean function for the continuous outcome variable *y* is generated under four settings: linear main effect model (LM), linear main and interaction effects model (LMI), nonlinear main effect model (NM) and nonlinear main and interaction effect model (NMI). In the settings of LMI and NMI, the 10 pairwise interactions of the five true features are also associated with the outcomes. For each of the four settings, we consider following four scenarios: sample size *n* = 1,000, *p* = 20 and *R*^2^ = 0.2, 0.1; and sample size *n* = 2,000, *p* = 40 and *R*^2^ = 0.2, 0.1. For the binary outcomes, we also generate data from four settings: logit link main effect model (Logit), logit link main and interaction effect model (LogitI), logit link nonlinear main effect model (Nlogit), and logit link nonlinear main and interaction effect model (NlogitI).

### Selection/Identification of Important Exposures in the Mixture

For continuous outcomes (Table 1: main/marginal), G-Lasso-MI has the lowest specificity (0.000) and highest false discovery rate (FDR=0.75) across all models, indicating it selects all 20 exposures. G-Lasso will either select a group of correlated predictors or shrink the whole group to zero. In our simulation settings, each group has a true predictor; G-Lasso hence selects all the exposures. Comparing Lasso-M and Lasso-MI or Enet-M and Enet-MI under all data settings, the inclusions of interactions into the models consistently decrease the sensitivity slightly (e.g., 0.975 to 0.954 in LM for lasso), increase the specificity (0.643 to 0.775) and decrease the FDR (0.497 to 0.391). BKMR shows low specificities in LM and LMI (0.082, 0.052), but high specificities in NM and NMI (0.913, 0.769). Comparing the three methods designed for interactions, HierNet demonstrates the highest sensitivity and highest FDR, while SNIF has the highest specificity and lowest FDR across all settings.

**Table 1:** Selection accuracy for main and interaction identification among eight methods, where continuous outcome is generated from LM, NM, LMI, and NMI. Means of Sen, Spe, FDR and FPR are obtained from 500 data replications with *N*_*train*_ = 500, *p* = 20, *q* = 5, and *R*^2^ = 0. 2

| Data | Type | Metric | Lasso-M | Enet-M | G-Lasso-M | Lasso-MI | Enet-MI | G-Lasso-MI | BKMR | RF | HigLasso | HierNet | SNIF |
| --- | --- | --- | --- | --- | --- | --- | --- | --- | --- | --- | --- | --- | --- |
| LM | Main/Marginal | Sen | 0.975 | 0.980 | 1.000 | 0.954 | 0.966 | 1.000 | 0.986 | 0.635 | 0.766 | 0.971 | 0.714 |
|  |  | Spe | 0.643 | 0.602 | 0.329 | 0.775 | 0.728 | 0.000 | 0.082 | 0.906 | 0.831 | 0.603 | 0.965 |
|  |  | FDR | 0.497 | 0.527 | 0.618 | 0.391 | 0.437 | 0.750 | 0.716 | 0.232 | 0.291 | 0.523 | 0.112 |
|  | Interaction | FPR | -- | -- | -- | 0.045 | 0.052 | 1.000 | -- | -- | 0.002 | 0.060 | 0.000 |
| LMI | Main/Marginal | Sen | 0.680 | 0.704 | 0.980 | 0.676 | 0.698 | 1.000 | 0.996 | 0.636 | 0.554 | 0.986 | 0.387 |
|  |  | Spe | 0.703 | 0.682 | 0.387 | 0.775 | 0.753 | 0.000 | 0.052 | 0.852 | 0.885 | 0.176 | 0.943 |
|  |  | FDR | 0.535 | 0.551 | 0.594 | 0.480 | 0.497 | 0.750 | 0.729 | 0.342 | 0.308 | 0.706 | 0.256 |
|  | Interaction | Sen | -- | -- | -- | 0.624 | 0.661 | 1.000 | -- | -- | 0.162 | 0.496 | 0.088 |
|  |  | Spe | -- | -- | -- | 0.894 | 0.885 | 0.000 | -- | -- | 0.992 | 0.907 | 0.999 |
|  |  | FDR | -- | -- | -- | 0.736 | 0.745 | 0.947 | -- | -- | 0.366 | 0.757 | 0.168 |
| NM | Main/Marginal | Sen | 0.614 | 0.634 | 1.000 | 0.552 | 0.571 | 1.000 | 0.588 | 0.421 | 0.664 | 0.902 | 0.597 |
|  |  | Spe | 0.733 | 0.692 | 0.316 | 0.820 | 0.783 | 0.000 | 0.913 | 0.950 | 0.928 | 0.514 | 0.979 |
|  |  | FDR | 0.516 | 0.559 | 0.632 | 0.447 | 0.493 | 0.750 | 0.145 | 0.153 | 0.151 | 0.594 | 0.071 |
|  | Interaction | FPR | -- | -- | -- | 0.093 | 0.105 | 1.000 | -- | -- | 0.002 | 0.062 | 0.000 |
| NMI | Main/Marginal | Sen | 0.649 | 0.670 | 1.000 | 0.588 | 0.611 | 1.000 | 0.732 | 0.474 | 0.684 | 0.944 | 0.620 |
|  |  | Spe | 0.712 | 0.682 | 0.298 | 0.800 | 0.771 | 0.000 | 0.769 | 0.921 | 0.931 | 0.464 | 0.976 |
|  |  | FDR | 0.532 | 0.554 | 0.642 | 0.472 | 0.502 | 0.750 | 0.285 | 0.215 | 0.149 | 0.611 | 0.080 |
|  | Interaction | Sen | -- | -- | -- | 0.288 | 0.310 | 1.000 | -- | -- | 0.086 | 0.227 | 0.011 |
|  |  | Spe | -- | -- | -- | 0.905 | 0.895 | 0.000 | -- | -- | 0.999 | 0.935 | 1.000 |
|  |  | FDR | -- | -- | -- | 0.845 | 0.851 | 0.947 | -- | -- | 0.040 | 0.826 | 0.016 |

*LM* linear main effects, *LMI* linear main effects and interactions, *NM* nonlinear main effects, *NMI* nonlinear main effects and interactions
*Sen* sensitivity, *Spe* specificity, *FDR* false discovery rate, *FPR* false positive rate

For binary outcomes (Table 2: main/marginal), similar to the results for continuous outcomes, G-Lasso-M and G-Lasso-MI have shown very low specificity and highest FDR. Comparing Lasso-M and Lasso-MI or Enet-M and Enet-MI under all data settings, we see similar trends as continuous outcomes, where sensitivity decreases, specificity increases, and FDR decreases after including interactions.

**Table 2:** Selection accuracy for main and interaction identification among five methods, where binary outcome is generated from Logit, LogitI, NLogitI, and NLogitI. Means of Sen, Spe, FDR and FPR are obtained from 500 data replications with *N*_*train*_ = 500, *p* = 20, *q* = 5 and *R*^2^ = 0. 2

| Data | Type | Metric | Lasso-M | Enet-M | G-Lasso-M | Lasso-MI | Enet-MI | G-Lasso-MI | RF | HierNet |
| --- | --- | --- | --- | --- | --- | --- | --- | --- | --- | --- |
| Logit | Main/Marginal | Sen | 0.965 | 0.978 | 0.980 | 0.919 | 0.963 | 1.000 | 0.557 | 0.908 |
|  |  | Spe | 0.635 | 0.543 | 0.293 | 0.803 | 0.691 | 0.000 | 0.752 | 0.589 |
|  |  | FDR | 0.507 | 0.567 | 0.624 | 0.367 | 0.470 | 0.750 | 0.523 | 0.498 |
|  | Interaction | FPR | -- | -- | -- | 0.057 | 0.087 | 1.000 | -- | 0.066 |
| LogitI | Main/Marginal | Sen | 0.930 | 0.954 | 0.988 | 0.822 | 0.892 | 0.996 | 0.515 | 0.920 |
|  |  | Spe | 0.650 | 0.558 | 0.177 | 0.816 | 0.727 | 0.004 | 0.706 | 0.520 |
|  |  | FDR | 0.504 | 0.563 | 0.679 | 0.377 | 0.461 | 0.747 | 0.594 | 0.558 |
|  | Interaction | Sen | -- | -- | -- | 0.260 | 0.343 | 0.996 | -- | 0.279 |
|  |  | Spe | -- | -- | -- | 0.937 | 0.909 | 0.004 | -- | 0.934 |
|  |  | FDR | -- | -- | -- | 0.800 | 0.816 | 0.944 | -- | 0.759 |
| Nlogit | Main/Marginal | Sen | 0.630 | 0.691 | 0.958 | 0.528 | 0.579 | 0.976 | 0.405 | 0.780 |
|  |  | Spe | 0.734 | 0.624 | 0.220 | 0.866 | 0.773 | 0.024 | 0.791 | 0.605 |
|  |  | FDR | 0.515 | 0.597 | 0.666 | 0.383 | 0.512 | 0.732 | 0.500 | 0.518 |
|  | Interaction | FPR | -- | -- | -- | 0.063 | 0.094 | 0.976 | -- | 0.053 |
| NlogitI | Main/Marginal | Sen | 0.717 | 0.772 | 0.954 | 0.578 | 0.648 | 0.962 | 0.410 | 0.846 |
|  |  | Spe | 0.712 | 0.599 | 0.236 | 0.863 | 0.759 | 0.038 | 0.770 | 0.560 |
|  |  | FDR | 0.512 | 0.590 | 0.656 | 0.373 | 0.502 | 0.722 | 0.530 | 0.530 |
|  | Interaction | Sen | -- | -- | -- | 0.183 | 0.251 | 0.962 | -- | 0.159 |
|  |  | Spe | -- | -- | -- | 0.937 | 0.901 | 0.038 | -- | 0.943 |
|  |  | FDR | -- | -- | -- | 0.851 | 0.867 | 0.911 | -- | 0.783 |

*Logit* logit-link linear main effects, *LogitI* logit-link linear main effects and interactions, *Nlogit* logit-link nonlinear main effects, *NlogitI* logit-link nonlinear main effects and interactions
*Sen* sensitivity, *Spe* specificity, *FDR* false discovery rate, *FPR* false positive rate
Mean prevalence of outcome over 500 replicates equals to 13.6%, 12.9%, 14.5% and 11.2% for Logit, LogitI, Nlogit and NlogitI, respectively

Excluding Glasso, Enet-M has the highest sensitivity in Logit and LogitI (0.978, 0.954), and HierNet has the highest sensitivity in Nlogit and NlogitI (0.780, 0.846). Lasso-MI demonstrates the highest specificity and the lowest FDR in all four settings (e.g., 0.803 and 0.367 in Logit).

### Interaction detection

For continuous outcomes (Table 1: interaction), G-Lasso-MI again exhibits a specificity of zero under LMI and NMI. For the remaining five methods, SNIF achieves the lowest false positive rate (FPR) of 0.000 in both LM and NM, while HierNet and Enet-MI have the highest FPR in LM (0.060) and NM (0.105), respectively. In LMI and NMI, Enet-MI demonstrates the highest sensitivity (0.661, 0.310), and SNIF again shows the highest specificity (0.999, 1.000) and lowest FDR (0.168, 0.016).

For binary outcomes (Table 2: interaction), G-Lasso/G-Lasso-MI have high FPR and low specificity. Lasso-MI and HierNet have the lowest FPR in Logit and Nlogit (0.057, 0.053). In LogitI and NlogitI, Enet-MI has the highest sensitivity (0.343, 0.251) and FDR (0.816, 0.867) among the three methods, while HierNet has slightly higher specificity and lower FDR. Overall, Lasso-MI, Enet-MI and HierNet produce similar results in interaction selection. However, the sensitivity for interactions in LogitI and NlogitI is low for all three methods, suggesting that identification of interactions is challenging for binary outcomes.

### Prediction of health outcome and risk stratification

For continuous outcomes (Table 3 “continuous outcome and continuous ERS/WQS/Q-gcomp”), in LM setting, Enet-M shows the highest correlation coefficient (Corr=0.43) and the smallest sum of squared errors (SSE=36.8), followed by SL with Corr of 0.42 and SSE of 37.2. In LMI, Enet-MI and SL have competitive Corr (0.39, 0.39) and SSE (155.4, 155.7). Since the true model LMI includes the interactions, models that account for interactions in general fit the data much better than models with only main effects. For instance, the Corr for Enet-M and Enet-MI are 0.19 and 0.39, respectively. This emphasizes the importance of including the interactions in the model fitting when there are true interactive associations. BKMR performs the worst among the five ERSs that account for interactions in terms of lowest Corr of 0.26. In NM, SL has the highest Corr of 0.40 and lowest SSE of 69.4. Note that even though the true model NM only has main effects, the ERS models considering interactions outperform the models considering only main effects. This is not surprising as interactions can capture nonlinear associations partially. Similar results are found in the NMI setting, where SL fits the data best and models considering interactions demonstrate advantages over models with only main effects. Additionally, it is worth mentioning that BKMR seems to fit nonlinear models better than linear models. When comparing WQS-M* with WQS-M or Q-gcomp-M* with Q-gcomp-M under, the models with variable selection outperform the models without variable selection under most data scenarios. Similar results are seen when comparing WQS-MI* with WQS-MI, where WQS-MI* has shown better or very similar Corr and SSE with WQS-MI. However, Q-gcomp-MI has shown a significant reduction in Corr and a large increase in SSE, suggesting that Q-gcomp-MI does not fit the full data well. It is also noteworthy that WQS and Q-gcomp models have shown similar results compared to the other methods under the LM setting, but their predictions are worse under other settings with interactions and/or nonlinearity.

**Table 3:** Risk prediction performance by different statistical methods, when data are generated from LM, NM, LMI, and NMI. Means of Corr, SSE, AUC, and median of OR are obtained from 500 data replications for *N*_*test*_ = 500, *p* = 20, *q* = 5, and *R*^2^ = 0. 2

| Data | Metric | ERS<br>Enet-M | WQS<br>-M* | WQS<br>-M | Q-gcomp<br>-M* | Q-gcomp<br>-M | ERS<br>Enet-MI | ERS<br>BKMR | ERS<br>HierNet | ERS<br>SNIF | ERS<br>SL | WQS<br>-MI* | WQS<br>-MI | Q-gcomp<br>-MI* | Q-gcomp<br>-MI |
| --- | --- | --- | --- | --- | --- | --- | --- | --- | --- | --- | --- | --- | --- | --- | --- |
| **Continuous Outcome and Continuous ERS/WQS/Q-gcomp** |  |  |  |  |  |  |  |  |  |  |  |  |  |  |  |
| LM | Corr | 0.43 | 0.41 | 0.41 | 0.40 | 0.39 | 0.42 | 0.32 | 0.42 | 0.41 | 0.42 | 0.39 | 0.33 | 0.37 | 0.20 |
|  | SSE | 36.8 | 37.6 | 37.6 | 37.8 | 38.3 | 37.4 | 40.8 | 37.3 | 37.6 | 37.2 | 38.4 | 40.0 | 39.4 | 66.0 |
| LMI | Corr | 0.19 | 0.19 | 0.20 | 0.18 | 0.16 | 0.39 | 0.26 | 0.36 | 0.28 | 0.39 | 0.34 | 0.34 | 0.32 | 0.17 |
|  | SSE | 176.1 | 176.9 | 176.5 | 178.0 | 180.8 | 155.4 | 172.6 | 159.2 | 176.3 | 155.7 | 162.4 | 161.7 | 170.7 | 275.0 |
| NM | Corr | 0.31 | 0.28 | 0.28 | 0.27 | 0.26 | 0.37 | 0.38 | 0.40 | 0.37 | 0.40 | 0.30 | 0.29 | 0.28 | 0.14 |
|  | SSE | 74.7 | 76.1 | 76.2 | 76.6 | 77.8 | 71.8 | 70.6 | 69.5 | 80.5 | 69.4 | 75.3 | 76.0 | 78.6 | 128.9 |
| NMI | Corr | 0.30 | 0.28 | 0.27 | 0.27 | 0.25 | 0.37 | 0.37 | 0.39 | 0.35 | 0.39 | 0.31 | 0.31 | 0.29 | 0.15 |
|  | SSE | 111.7 | 113.6 | 113.9 | 114.3 | 116.2 | 106.4 | 106.4 | 104.4 | 121.9 | 104.3 | 111.4 | 111.5 | 116.1 | 189.5 |
| **Dichotomous Outcome and Continuous ERS/ WQS/Q-gcomp** |  |  |  |  |  |  |  |  |  |  |  |  |  |  |  |
| LM | AUC | 0.73 | 0.72 | 0.72 | 0.72 | 0.72 | 0.73 | 0.68 | 0.73 | 0.72 | 0.73 | 0.72 | 0.70 | 0.71 | 0.62 |
| LMI | AUC | 0.65 | 0.65 | 0.65 | 0.64 | 0.63 | 0.73 | 0.67 | 0.72 | 0.68 | 0.73 | 0.72 | 0.71 | 0.71 | 0.64 |
| NM | AUC | 0.70 | 0.68 | 0.68 | 0.68 | 0.67 | 0.72 | 0.73 | 0.74 | 0.73 | 0.74 | 0.69 | 0.68 | 0.68 | 0.60 |
| NMI | AUC | 0.70 | 0.69 | 0.69 | 0.68 | 0.67 | 0.72 | 0.72 | 0.73 | 0.72 | 0.73 | 0.70 | 0.69 | 0.69 | 0.61 |
| **Dichotomous Outcome and Categorical ERS/ WQS/Q-gcomp** |  |  |  |  |  |  |  |  |  |  |  |  |  |  |  |
| LM | OR | 11.2 | 10.0 | 9.8 | 9.8 | 9.4 | 10.4 | 8.0 | 10.8 | 10.3 | 10.8 | 9.0 | 6.1 | 8.2 | 3.0 |
| LMI | OR | 2.9 | 2.9 | 3.0 | 2.8 | 2.7 | 6.7 | 5.8 | 5.6 | 4.3 | 6.9 | 6.0 | 5.7 | 5.4 | 3.1 |
| NM | OR | 5.6 | 5.0 | 4.9 | 5.1 | 4.7 | 7.2 | 7.8 | 8.7 | 7.0 | 8.7 | 5.8 | 4.9 | 5.1 | 2.4 |
| NMI | OR | 5.5 | 5.0 | 4.9 | 4.9 | 4.7 | 7.2 | 6.8 | 7.4 | 6.3 | 7.8 | 5.5 | 5.1 | 5.3 | 2.6 |

*LM* linear main effects, *LMI* linear main effects and interactions, *NM* nonlinear main effects, *NMI* nonlinear main effects and interactions
*Corr* correlation, *SSE* sum of squared error, *AUC* area under the receiver operating characteristic curve, *OR* odds ratio
*Enet-M* elastic net for main effects, *WQS-M\** weighted quantile sum regression (WQS) for selected main effects by Enet-M, *WQS-M* WQS for main effects, *Q-gcomp-M\** quantile g-computation (Q-gcomp) for selected main effects by Enet-M, *Q-gcomp-M* Q-gcomp for main effects, *Enet-MI* elastic net for main effects and interactions, *BKMR* Bayesian kernel machine regression, *HierNet* lasso for hierarchical interactions, *SNIF* selection of nonlinear interactions by a forward stepwise algorithm, *SL* super learner, *WQS-MI\** WQS for selected main effects and interactions by Enet-MI, *WQS-MI* WQS for main effects and interactions, *Q-gcomp-MI\** Q-gcomp for selected main effects and interactions by Enet-MI, *Q-gcomp-MI* Q-gcomp for main effects and interactions

For binary outcomes dichotomized from continuous outcome with the continuous ERS/WQS/Q-gcomp, the area under the ROC curve (AUC) results are consistent with those of Corr and SSE. In LM, Enet-M, Enet-MI, HierNet and SL all achieve the highest AUC of 0.73; while in the remaining three settings, the five ERS methods considering interactions have a higher AUC than the methods with only main effects. SL achieves the highest AUC in all settings. For categorical ERS/WQS/Q-gcomp, Enet-M is top-performing with highest risk stratification odds ratio (OR=11.2) followed by HierNet and SL (OR=10.8) in LM, and SL has highest OR in LMI, NM and NMI. To summarize Table 3, SL is the top-performing method across all settings, demonstrating the strength of its ensemble algorithm that combines multiple learners.

For binary outcomes (Table 4), Enet-M has the highest AUC of 0.812 and lowest Brier of 0.096 and highest OR of 33.5 in Logit setting, followed by SL (AUC=0.801) and Lasso-MI, Enet-MI (Brier=0.098) and WQS-M* (OR=28.6). In LogitI, the three models that only consider main effects, have higher or similar AUC and higher OR than the five ERS methods that incorporate interactions. In Nlogit and NlogitI, HierNet and SL achieve the highest AUC (0.753 and 0.780) and high ORs (11.3, 14.1). For WQS and Q-gcomp, the results comparing the models with and without variable selection show that the models with variable selection outperform the full models in terms of higher AUC, lower or similar Brier score, and higher risk stratification OR regardless of settings. It is evident that variable selection can greatly improve the prediction accuracy for these two methods.

**Table 4:** Risk prediction performance by different statistical methods, when data are generated from Logit, LogitI, Nlogit, and NLogitI. Means of AUC and Brier, and median of OR are obtained from 500 data replications for *N*_*test*_ = 500, *p* = 20, *q* = 5 and *R*^2^ = 0. 2

| Data | Metric | ERS<br>Enet-M | WQS<br>-M* | WQS<br>-M | Q-gcomp<br>-M* | Q-gcomp<br>-M | ERS<br>Lasso-MI | ERS<br>Enet-MI | ERS<br>RF | ERS<br>HierNet | ERS<br>SL | WQS<br>-MI* | WQS<br>-MI | Q-gcomp<br>-MI* | Q-gcomp<br>-MI |
| --- | --- | --- | --- | --- | --- | --- | --- | --- | --- | --- | --- | --- | --- | --- | --- |
| **Dichotomous Outcome and Continuous ERS WQS/Q-gcomp** |  |  |  |  |  |  |  |  |  |  |  |  |  |  |  |
| Logit | AUC | 0.812 | 0.799 | 0.798 | 0.795 | 0.787 | 0.800 | 0.800 | 0.778 | 0.796 | 0.801 | 0.770 | 0.740 | 0.763 | 0.594 |
|  | Brier | 0.096 | 0.099 | 0.099 | 0.100 | 0.103 | 0.098 | 0.098 | 0.101 | 0.100 | 0.099 | 0.102 | 0.104 | 0.109 | 0.273 |
| LogitI | AUC | 0.771 | 0.762 | 0.761 | 0.758 | 0.749 | 0.756 | 0.757 | 0.735 | 0.761 | 0.759 | 0.732 | 0.721 | 0.723 | 0.589 |
|  | Brier | 0.094 | 0.096 | 0.097 | 0.097 | 0.099 | 0.094 | 0.094 | 0.098 | 0.095 | 0.094 | 0.097 | 0.098 | 0.104 | 0.271 |
| Nlogit | AUC | 0.751 | 0.733 | 0.731 | 0.727 | 0.714 | 0.750 | 0.748 | 0.732 | 0.753 | 0.753 | 0.712 | 0.698 | 0.702 | 0.561 |
|  | Brier | 0.108 | 0.112 | 0.112 | 0.114 | 0.116 | 0.109 | 0.109 | 0.110 | 0.109 | 0.108 | 0.114 | 0.114 | 0.121 | 0.297 |
| NlogitI | AUC | 0.778 | 0.760 | 0.759 | 0.755 | 0.743 | 0.779 | 0.776 | 0.762 | 0.780 | 0.780 | 0.742 | 0.727 | 0.729 | 0.586 |
|  | Brier | 0.085 | 0.088 | 0.088 | 0.089 | 0.092 | 0.084 | 0.085 | 0.086 | 0.085 | 0.085 | 0.089 | 0.089 | 0.097 | 0.254 |
| **Dichotomous Outcome and Categorical ERS WQS/Q-gcomp** |  |  |  |  |  |  |  |  |  |  |  |  |  |  |  |
| Logit | OR | 33.5 | 28.6 | 27.2 | 27.0 | 23.4 | 23.9 | 24.1 | 15.4 | 25.5 | 25.6 | 13.6 | 8.7 | 14.2 | 2.1 |
| LogitI | OR | 10.5 | 10.2 | 9.8 | 9.8 | 9.4 | 9.0 | 9.2 | 4.8 | 9.5 | 9.1 | 7.1 | 6.6 | 6.6 | 2.0 |
| Nlogit | OR | 11.0 | 9.6 | 9.0 | 9.3 | 8.1 | 10.7 | 10.5 | 6.7 | 11.3 | 11.2 | 7.3 | 6.0 | 6.6 | 1.6 |
| NlogitI | OR | 13.6 | 11.9 | 11.4 | 11.7 | 10.6 | 13.7 | 12.9 | 8.7 | 13.9 | 14.1 | 8.7 | 7.9 | 8.7 | 2.0 |

*Logit* logit-link linear main effects, *LogitI* logit-link linear main effects and interactions, *Nlogit* logit-link nonlinear main effects, *NlogitI* logit-link nonlinear main effects and interactions
*AUC* area under the receiver operating characteristic curve, *Brier* Brier score, *OR* odds ratio
*Enet-M* elastic net for main effects, *WQS-M\** weighted quantile sum regression (WQS) for selected main effects by Enet-M, *WQS-M* WQS for main effects, *Q-gcomp-M\** quantile g-computation (Q-gcomp) for selected main effects by Enet-M, *Q-gcomp-M* Q-gcomp for main effects, *Lasso-MI* lasso for main effects and interactions, *Enet-MI* elastic net for main effects and interactions, *RF* random forest, *HierNet* lasso for hierarchical interactions, *SL* super learner, *WQS-MI\** WQS for selected main effects and interactions by Enet-MI, *WQS-MI* WQS for main effects and interactions, *Q-gcomp-Mi\** Q-gcomp for selected main effects and interactions by Enet-MI, *Q-gcomp-MI* Q-gcomp for main effects and interactions
Mean $R^2$ over 500 replicates equals to 0.22, 0.22, 0.18 and 0.23 for Logit, LogitI, Nlogit and NlogitI, respectively
Mean prevalence of outcome over 500 replicates equals to 13.6%, 12.9%, 14.5% and 11.2% for Logit, LogitI, Nlogit and NlogitI, respectively

### Computing time

To compare the computing time for each method, Table S13 lists the mean computing time in seconds for each method under various data settings when signals are small. Specifically, we consider settings of *N*_*train*_= 500 and *p* = 20 or *N*_*train*_= 1,000 and *p* = 40 with *R*^2^ = 0.1 for continuous outcomes and *R*^2^ = 0.1 for binary outcomes. We run each setting with 100 data replications then calculate the mean time each takes. There is a significant difference in computing time among these methods, with Q-gcomp being the fastest, requiring as little as 0.03 seconds, followed by RF, Enet, Lasso, SNIF and WQS, all of which with negligible computing time, and BKMR (2000 MCMC iterations) and HigLasso being the slowest, requiring 2,000 to 50,000 seconds. The computing time varies only slightly for different true data settings for the same sample size/number of pollutants. However, computing time increases significantly when sample size increases from 500 to 1000 and number of pollutants increases from 20 to 40.

### Summary

Based on the simulation results presented in Tables 1-4 and S1-S12, we summarize recommendations for the methods in Table 5 under different settings of sample sizes, number of pollutants, and small or medium signals for continuous and binary outcomes. For continuous outcomes, the results consistently suggest that HierNet, SNIF and Enet-MI have the most stable selections for pollutants and their interactions. For pollutant selection, HierNet almost always shows the highest sensitivity, while SNIF exhibits the lowest sensitivity but the highest specificity and lowest FDR, suggesting even though it may miss important pollutants, it tends to select only the true significant pollutants. Depending on the specific research question, we recommend that researchers utilize both HierNet and SNIF to compare the results in the real data analysis, considering whether they prefer sensitivity or control of FDR. For interaction selection, Enet-MI and SNIF seem to perform better than the other methods, but in general, the sensitivity is very low and FDR is high. This suggests that there is a clear need for the development of new methods for detection of interactions. For prediction, SL and HierNet perform better than the rest of methods. SL has the advantage of ensemble learning from predictions via multiple methods, and HierNet, even though a linear method, can fit nonlinear data by selecting the interactions and quadratic terms.

**Table 5:** Recommendation table of the methods under various data settings.

| **Continuous outcome** | **Sample size medium (n=1000/p=20)** |  | **Sample size large (n=2000/p=40)** |  |
| --- | --- | --- | --- | --- |
|  | **Signal medium**<br>**Tables 2&4** | **Signal small**<br>**Tables S1&S2** | **Signal medium**<br>**Tables S5&S6** | **Signal small**<br>**Tables S9&S10** |
| Pollutant selection | HierNet, SNIF | HierNet, SNIF | HierNet, SNIF | HierNet, SNIF |
| Interaction detection | Enet-MI, SNIF | Enet-MI, SNIF | Enet-MI, SNIF | Enet-MI, SNIF |
| Prediction | SL, HierNet | SL, HierNet | SL, HierNet | SL, Enet-M |
| **Binary outcome** | **Sample size medium (n=1000/p=20)** |  | **Sample size large (n=2000/p=40)** |  |
|  | **Signal medium**<br>**Tables 3&5** | **Signal small**<br>**Tables S3&S4** | **Signal medium**<br>**Tables S7&S8** | **Signal small**<br>**Tables S11&S12** |
| Pollutant selection | Enet-M, Lasso-MI,<br>HierNet | Enet-M, Lasso-MI | Lasso-MI, HierNet | Enet-M, Lasso-MI,<br>HierNet |
| Interaction detection | Enet-MI, HierNet | Enet-MI, HierNet | Lasso-MI, Enet-MI | Enet-MI, HierNet |
| Prediction | Enet-M, SL | Enet-M | Enet-M, SL | Enet-M, SL |

For binary outcomes, there are limited methods available to use. For pollutant selection, Enet-M, Lasso-MI and HierNet exhibit satisfactory sensitivity, but their FDRs are relatively high. For interaction detection, G-lasso-MI has low specificity, so the options remaining are Enet-MI, Lasso-MI and HierNet. Unfortunately, these methods suffer from low sensitivity and high FDR, making the selection for interactions in binary outcomes quite challenging. For prediction, Enet-M outperforms many methods under various settings, suggesting that a parsimonious model might achieve the same or better prediction accuracy compared to other larger models for binary outcomes. For WQS and Q-gcomp, the results show that the models with variable selection provide higher AUC, lower or similar Brier score, and higher risk stratification OR regardless of settings than the models without variable selection.

## PROTECT Data Analysis Results

### Important pollutants, covariates, and interactions

Table 6 reports the variables that are selected at least 30% of the time by each method in the 100 fittings using random training data. For birth weight, Enet-M selects two metals (Ba and As) and one phthalate (MCOP), and Figures S1 in the Supplementary Material show the distributions of the 100 coefficient estimates for these three chemicals, indicating positive associations with birth weight when Ba is selected and negative associations when As and MCOP are selected. Note that Enet-M can only select 39 chemicals as covariates are controlled. Enet-MI only selects one main effect “gestational age”, and it tends to select interactions over main effects compared with Enet-M. BKMR selects all 39 chemicals at least 30 times, so we report the top eight compounds that are most frequently selected, ranging from 47 to 55 times out of 100 times. The frequently selected chemicals are three metals (Ba, As and Sn), two phthalates (MBZP and MCOP), two phenols (BP3 and TCS), and one PAH (4PHE). HierNet selects 16 main effects, of which seven are metals, and nine interactions, eight of which involve gestational age and a chemical. SNIF only selects main effects of metals and covariates, and based on the simulations, SNIF tends to be conservative in selection as evidenced by the lowest FDR, indicating that the exposures or covariates it selects are almost always true predictors. Comparing the selections across methods, we find that metals (Ba, As and Co), phthalates (MBZP and MCOP), and phenol (BPA) are frequently selected as main effects or interactions.

**Table 6:**
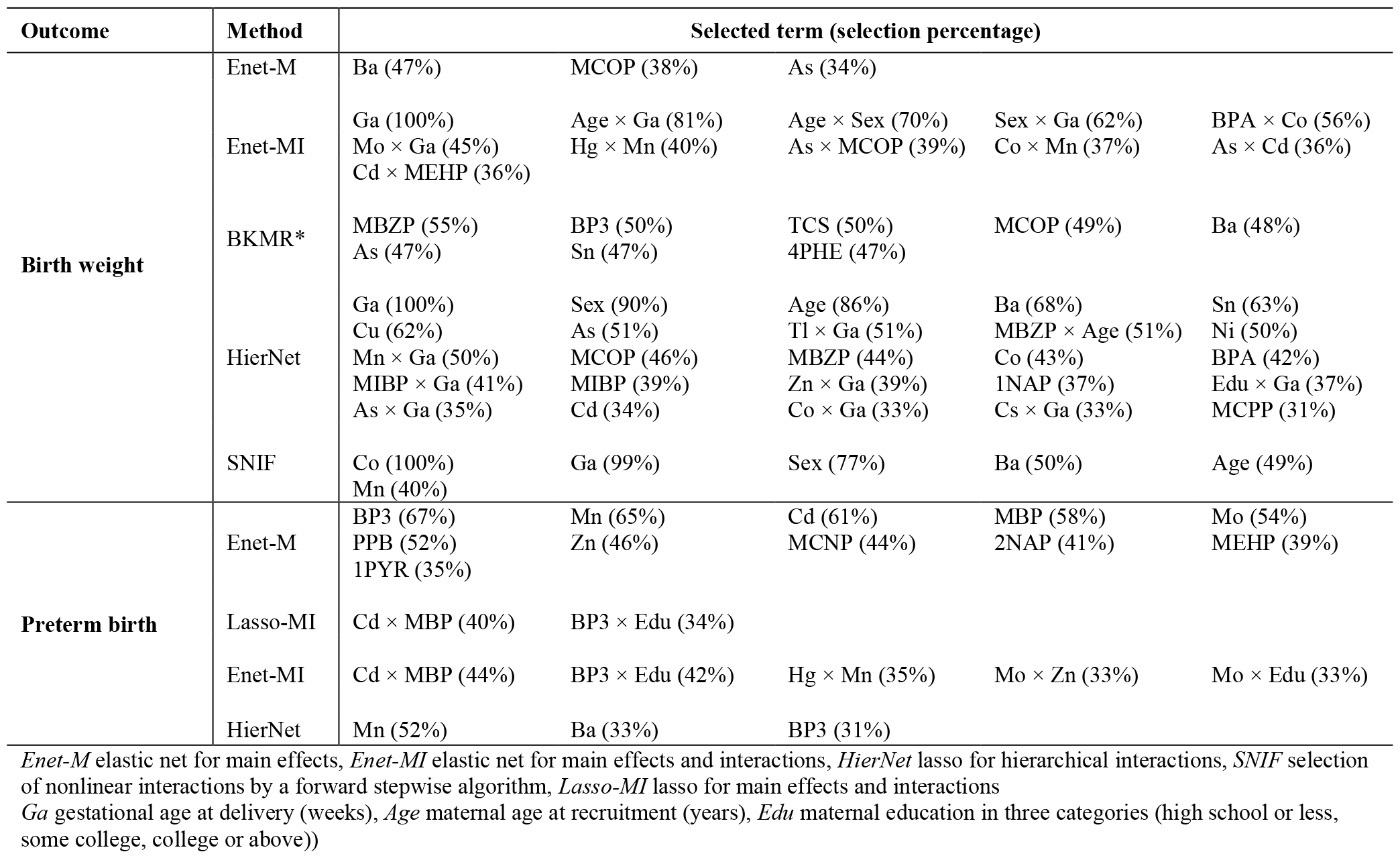
Main and interaction effects selected at least 30% of the 100 random sampled training data.

For preterm birth, we do not report the variable selection by RF as it is not designed for this purpose. Lasso-MI and Enet-MI tend to select interactions over main effects, whereas HierNet selects main effects only. The metals Mn and Cd or their interactions with other chemicals are selected across methods. The only covariate selected is maternal education. It is worth noting that for both birth weight and preterm outcomes, HierNet also screens for any quadratic terms, but we omit summarizing them in this analysis as quadratic term selection is not a focus of this paper.

### Predictive Power Comparison

Table 7 reports a comparison of the predictive power among summary scores of ERS, WQS and Q-gcomp for different outcomes. For birth weight, the mean weight for SL across 100 times of random splits for Enet-MI, BKMR, HierNet and SNIF are 55.3%, 1.0%, 35.1% and 8.7%, indicating that Enet and HierNet have better overall predictive performance. Enet-M and SL outperform other methods in terms of Corr and SSE for main effect and main and interaction effect models, respectively. For the low birth weight binary outcome, HierNet and SL achieve highest AUC (0.838) and the highest ORs of having low birth weight (12.73, 12.53) when comparing the lowest quartiles of ERSs versus the rest of the samples. For the high birth weight binary outcome, WQS-M and Enet-M achieve higher AUC (0.665, 0.664) than the other methods, suggesting that main effect models fit the data better for the high birth weight outcome. WQS-M also yields the highest OR of having high birth weight (2.70) when comparing the highest quartiles of predictive values versus the rest of the samples.

**Table 7:** Comparison of risk prediction performance by different methods.

| Outcome |  | ERS<sub>Enet-M</sub> | WQS-M | Q-gcomp-M | ERS<sub>Enet-MI</sub> | ERS<sub>BKMR</sub> | ERS<sub>HierNet</sub> | ERS<sub>SNIF</sub> | ERS<sub>SL</sub> |
| --- | --- | --- | --- | --- | --- | --- | --- | --- | --- |
|  |  | Continuous birth weight and continuous ERS |  |  |  |  |  |  |  |
| Birth weight | Corr | 0.542 | 0.542 | 0.510 | 0.533 | 0.512 | 0.522 | 0.479 | 0.534 |
|  | SSE | 0.197 | 0.198 | 0.208 | 0.200 | 0.208 | 0.204 | 0.310 | 0.200 |
|  | Low birth weight and continuous ERS |  |  |  |  |  |  |  |  |
|  | AUC | 0.835 | 0.836 | 0.826 | 0.837 | 0.830 | 0.838 | 0.822 | 0.838 |
|  | Low birth weight and categorical ERS (Q<sub>1</sub> vs rest) |  |  |  |  |  |  |  |  |
|  | OR | 11.82 | 12.11 | 9.97 | 12.49 | 11.22 | 12.73 | 11.79 | 12.53 |
|  | High birth weight and continuous ERS |  |  |  |  |  |  |  |  |
|  | AUC | 0.665 | 0.664 | 0.631 | 0.657 | 0.652 | 0.657 | 0.629 | 0.658 |
|  |  | High birth weight and categorical ERS (Q<sub>4</sub> vs rest) |  |  |  |  |  |  |  |
|  | OR | 2.52 | 2.70 | 1.98 | 2.33 | 2.43 | 2.56 | 2.10 | 2.43 |
|  |  | ERS<sub>Enet-M</sub> | WQS-M | Q-gcomp-M | ERS<sub>Lasso-MI</sub> | ERS<sub>Enet-MI</sub> | ERS<sub>RF</sub> | ERS<sub>HierNet</sub> | ERS<sub>SL</sub> |
|  |  | Preterm birth and continuous ERS |  |  |  |  |  |  |  |
| Preterm birth | AUC | 0.597 | 0.570 | 0.594 | 0.547 | 0.544 | 0.549 | 0.527 | 0.553 |
|  | Brier | 0.083 | 0.083 | 0.088 | 0.083 | 0.083 | 0.086 | 0.083 | 0.083 |
|  | Preterm birth and ERS (high vs low risk) |  |  |  |  |  |  |  |  |
|  | OR | 2.31 | 1.91 | 2.31 | 1.56 | 1.49 | 2.00 | 1.23 | 1.73 |

The covariates accounted for in birth weight are infant sex, gestational age at delivery (weeks), education (high school or less, some college, college or above) and maternal age at recruitment (years); covariates accounted for preterm include aforementioned variables except gestational age. The covariates are adjusted and not penalized in models of Enet-M, WQS-M, Q-gcomp-M, and BKMR; while covariates can be selected in Enet-MI, HierNet and SNIF.
*Corr* correlation, *SSE* sum of squared error, *AUC* area under the receiver operating characteristic curve, *OR* odds ratio, *Brier* Brier score *Enet-M* elastic net for main effects, *WQS-M* weighted quantile sum regression (WQS) for main effects, *Q-gcomp-M* quantile g-computation (Q-gcomp) for main effects, *Enet-MI* elastic net for main effects and interactions, *BKMR* Bayesian kernel machine regression, *HierNet* lasso for hierarchical interactions, *SNIF* selection of nonlinear interactions by a forward stepwise algorithm, *SL* super learner, *Lasso-MI* lasso for main effects and interactions, *RF* random forest

**Table 8.** Complete list of simulation settings.

| Model | Outcome | Sample size | Number of pollutants | Number of true effects | $R^2$ | Mean function* |
| --- | --- | --- | --- | --- | --- | --- |
| LM | continuous | 1000 | 20 | 5 | 0.2 | (1) |
| LMI | continuous | 1000 | 20 | 5 | 0.2 | (2) |
| NM | continuous | 1000 | 20 | 5 | 0.2 | (3) |
| NMI | continuous | 1000 | 20 | 5 | 0.2 | (4) |
| Logit | binary | 1000 | 20 | 5 | 0.2 | (1) |
| LogitI | binary | 1000 | 20 | 5 | 0.2 | (2) |
| Nlogit | binary | 1000 | 20 | 5 | 0.2 | (3) |
| NlogitI | binary | 1000 | 20 | 5 | 0.2 | (4) |
| LM | continuous | 2000 | 40 | 5 | 0.2 | (1) |
| LMI | continuous | 2000 | 40 | 5 | 0.2 | (2) |
| NM | continuous | 2000 | 40 | 5 | 0.2 | (3) |
| NMI | continuous | 2000 | 40 | 5 | 0.2 | (4) |
| Logit | binary | 2000 | 40 | 5 | 0.2 | (1) |
| LogitI | binary | 2000 | 40 | 5 | 0.2 | (2) |
| Nlogit | binary | 2000 | 40 | 5 | 0.2 | (3) |
| NlogitI | binary | 2000 | 40 | 5 | 0.2 | (4) |
| LM | continuous | 1000 | 20 | 5 | 0.1 | (1) |
| LMI | continuous | 1000 | 20 | 5 | 0.1 | (2) |
| NM | continuous | 1000 | 20 | 5 | 0.1 | (3) |
| NMI | continuous | 1000 | 20 | 5 | 0.1 | (4) |
| Logit | binary | 1000 | 20 | 5 | 0.1 | (1) |
| LogitI | binary | 1000 | 20 | 5 | 0.1 | (2) |
| Nlogit | binary | 1000 | 20 | 5 | 0.1 | (3) |
| NlogitI | binary | 1000 | 20 | 5 | 0.1 | (4) |
| LM | continuous | 2000 | 40 | 5 | 0.1 | (1) |
| LMI | continuous | 2000 | 40 | 5 | 0.1 | (2) |
| NM | continuous | 2000 | 40 | 5 | 0.1 | (3) |
| NMI | continuous | 2000 | 40 | 5 | 0.1 | (4) |
| Logit | binary | 2000 | 40 | 5 | 0.1 | (1) |
| LogitI | binary | 2000 | 40 | 5 | 0.1 | (2) |
| Nlogit | binary | 2000 | 40 | 5 | 0.1 | (3) |
| NlogitI | binary | 2000 | 40 | 5 | 0.1 | (4) |

\*Mean functions
(1) $x_1 + x_2 + x_3 + x_4 + x_5$
(2) $x_1 + x_2 + x_3 + x_4 + x_5 + x_1x_2 + x_1x_3 + x_1x_4 + x_1x_5 + x_2x_3 + x_2x_4 + x_2x_5 + x_3x_4 + x_3x_5 + x_4x_5$
(3) $x_1I(x_1 > 0) + \exp(x_2) + |x_3| + x_4^2 + (x_5 + 1)^2$
(4) $x_1I(x_1 > 0) + \exp(x_2) + |x_3| + x_4^2 + (x_5 + 1)^2 + x_1 \exp(x_2)I(x_1 > 0) + x_1|x_3|I(x_1 > 0) + x_1x_4^2I(x_1 > 0) + x_1(x_5 + 1)^2I(x_1 > 0) + \exp(x_2)|x_3| + \exp(x_2)x_4^2 + \exp(x_2)(x_5 + 1)^2 + |x_3|x_4^2 + |x_3|(x_5 + 1)^2 + x_4^2(x_5 + 1)^2$
*LM* linear main effects, *Logit* logit link linear main effects, *LMI* linear main effects and interactions, *LogitI* logit link linear main effects and interactions, *NM* nonlinear main effects, *Nlogit* logit link nonlinear main effects, *NMI* nonlinear main effects and interactions, *NlogitI* logit link nonlinear main effects and interactions

For preterm binary outcome, the mean weight for SL across 100 times of random splits for Lasso, Enet, RF and HierNet are 20.8%, 11.1%, 10.8% and 57.3%, respectively, indicating that HierNet has better overall predictions than the other three approaches. The main effect models give higher AUC than the four individual ERSs accounting for interactions, where ERS-M has the highest AUC (0.597). Enet-M, WQS-M, Enet-MI, BKMR and SL all achieve the smallest Brier scores (0.083). ERS-M and Q-gcomp-M have the highest OR of having a preterm birth when comparing the highest and lowest quartiles of summary measures. For the main and interaction models, SL has the best AUC, Brier and HierNet has the highest OR.

## Software

To facilitate the implementation of the statistical methods among practitioners, we have developed an open-source R package “CompMix: A comprehensive toolkit for environmental mixtures analysis”, currently featuring the implementation of 8 methods, including Lasso, Enet, BKMR, RF, HierNet, SNIF, WQS and Q-gcomp for continuous outcomes, and 6 methods, including Lasso, Enet, RF, HierNet, WQS and Q-gcomp for binary outcomes. The package offers the flexibility to perform three tasks: (1) toxicant selection and (2) interaction detection under various circumstances, and (3) the prediction performance across different models for users to determine which models fit their data best. Our package offers several unique features to existing software: first, it provides easy-used interface with few input arguments. All tuning parameters have been set default values tested by extensive simulation studies, greatly facilitating off-the-shelf tuning parameter selection. On the other hand, the package also provides an interface to modify the tuning parameters and model specifications for statisticians who are more familiar with those existing packages. Second, the users can also select one specific method, and examine results from different model specifications. For example, if the user would like to implement Lasso, the package will carry out the data analysis with different options, such as whether or not to perform the selection on interactions between exposures and/or covariates. Lastly, this package also provides a comprehensive summary of model fit, which offers useful information for the users to select the appropriate methods for their data. We will update this software regularly and include more emerging methods as they become available in the future. The package can be downloaded from the Comprehensive R Archive Network.

## Discussion

This paper presents an analytic framework to study the association between exposure to chemical mixtures and health outcomes. We evaluate several statistical methods for three research questions in mixtures analyses through simulation studies that range from simple linear models to complex nonlinear models. While the methods evaluated in this paper are not exhaustive, they represent a diverse set of approaches with unique strengths that can be utilized to answer specific research questions. To enhance the prediction accuracy among ERSs, we propose a method inspired by SL, where we iteratively solve weights for each candidate learner and combine their predictions using their weighted sum of ERSs. We have developed an R package “CompMix: A comprehensive toolkit for environmental mixtures analysis” for practitioners to analyze their data and compare results across versatile methods.

**Lessons learned from simulation studies:** our simulation studies for continuous outcomes demonstrate that for pollutant selection, HierNet almost always shows the highest sensitivity; for interaction detection, Enet-MI and SNIF seem to perform better than the other methods; for prediction, SL and HierNet outperform other methods across the settings, highlighting SL’s strength as an ensemble algorithm that combines multiple learners. For pollutant and interaction selection with a binary outcome, all the investigated methods either exhibit high sensitivity and high FDR, or low sensitivity. For prediction, Enet-M outperforms many methods under various settings, suggesting that a parsimonious model might achieve the same or better prediction accuracy compared to other larger models.

Furthermore, we notice that regardless of whether the true data are generated with interactions or not, fitting models that account for nonlinearity (such as BKMR) or include interactions generally yield better results than models with only main effects. Thus, we recommend considering models that accommodate interaction and nonlinearity in addition to linear models.

**New insights from PROTECT data analysis:** metals (Ba, As and Co), phthalates (MBZP, MCOP), and phenol (BPA) are more likely to be associated with the birth weight after adjusting for possible confounding factors such as such as age, gestational age at delivery. In particular, the interaction effects between Co and BPA on birth weight are more frequently identified compared with others. Our analysis also indicates that metals Mn and Cd and their interactions may have high impact on the preterm birth. All these findings are confirmed by different methods, which deserve further investigations by environmental epidemiologists.

**Limitations of the current study:** first, our simulation studies did not include any covariates in the model when comparing different methods. This is because some methods such as HierNet and SNIF cannot separate the covariates from pollutants for selection. However, in real world data analysis, the associations between the outcome and the predictors are much more complex, requiring the inclusion of multiple categorical and continuous covariates. Second, our simulation studies only focus on dataset with complete observations, while some chemicals in the PROTECT study may involve the high percentages of measurements below limit of detection (LOD) or missing measurements across three visits. The impact of imputation methods on the data analysis and scientific findings are worth further investigating. Third, our analysis for real data in the PROTECT study did not include all possible confounders such as family income, parity, occupation, and others, which may influence both exposures and outcome.

**Open problems and future directions:** to further study the health impact of mixtures, new and efficient statistical methods are urgently needed to address many important issues, including missing data and measurement errors [29], longitudinal measurements [30], nonlinear interaction detection [31], integrating multi-omics data [32], mediation analysis with mixtures [33, 34], causal inference with mixtures [35]. Estimating the health effect from exposure to chemical mixtures is a complex and challenging topic that requires a multidisciplinary team comprising epidemiologists, statisticians, and toxicologists. This team must work together to formulate the scientific question, identify the statistical barriers, interpret the study findings, and understand the limitations of the research. Through close collaborations, innovative methods can be designed and implemented in mixtures research to enhance our understanding of the health impacts from exposure to chemical mixtures.

## Methods

We will provide a detailed overview of current statistical methods for mixtures analysis, with a focus on supervised methods that can be implemented to construct ERS. To begin, we introduce the notations and problem setup. Consider a random sample of *N* subjects. For subject *i*(*i* = 1, …, *N*), let *x*_*i,p*_ denote the *p*th environmental pollutant exposure, *p* = 1, …, *P*; *z*_*i,k*_ denote the *k*th confounding factor, *k* = 1, …, *K*; *y*_*i*_ denote the one dimensional continuous or binary outcome of interest. Let 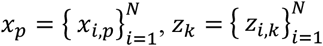 and 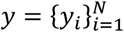. Let *D*_i_ = (*x*_*i*,1_, … *x*_*i,P*_, *z*_*i*,1_, …, *z* _*i,K*_, *y*_*i*_) represent the observed data for subject *i*. Additionally, we define 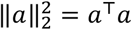 for any vector *a*. For simplicity of the illustration and without loss of generality, we assume that confounders are not present when reviewing some existing literature.

### Group 1 Methods

This group of approaches aim to perform the variable selection of the main effects of the pollutants. They can be divided into two categories: penalized regression approaches and machine learning approaches.

#### Lasso

The Least absolute shrinkage and selection operator (Lasso) was proposed by Tibshirani in 1996 [10]. It is a broadly used linear regression method that performs feature selection by penalizing the sum of the absolute values of the coefficients, termed as L_1_ penalty. It was developed to improve the prediction accuracy by selecting the most important predictors, while shrinking other coefficients to zero. Lasso minimizes the following objective function,

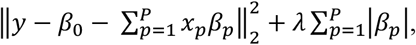

where *β*_*p*_(*p* = 0, …, *P*) are model parameters, and *λ* (*λ* ≥ 0) is the tuning parameter to regulate the size of parameters that are shrunk to zero. When *λ* = 0, Lasso is equivalent to ordinary linear regression, as *λ* increases, many regression coefficients *β*_*p*_ are shrunk to zero. In our analysis, we select the *λ* value that gives the minimum mean cross-validated error. One limitation of Lasso is that when handling a group correlated predictors, it often selects one pollutant from the group while shrinking the coefficients of other members to zero. We implement Lasso via R package “glmnet” (version 4.1-4) [36].

#### Enet

The Elastic net (Enet) was proposed by Zou et al. in 2005 [11]. It is also a variable selection and penalized regression method and it addresses the issue of L_1_ penalty that Lasso used by adding an L_2_ penalty, which is the sum of the squared coefficients. This advantage of Elastic net is especially appealing in the context of mixture analysis, where exposure to multipollutant within the same family class tend to be highly correlated. Enet minimizes the following objective function,

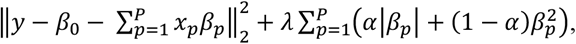

where 0 ≤ *α* ≤ 1 is another tunning parameter in addition to *λ* and *α* controls weights between L_1_ and L_2_ penalties. In our analysis, we specify *α* = 0.5 and select *λ* value that gives the minimum mean cross-validated error. We implement Enet via R package “glmnet” (version 4.1-4) [36].

#### G-Lasso

Group Lasso (G-Lasso) was proposed by Yuan et al. in 2006 [12]. It is an extension of Lasso that performs variable selections on prespecified groups of variables. It allows the entire group of variables to be either included or excluded from the model. As Enet, this unique feature is particularly suitable since multipollutant exposure are often correlated and presented in groups. However, not all pollutants in a group may be relevant to the outcome, despite being highly correlated. Therefore, G-Lasso may potentially have a high FDR. Suppose the data consists of *G* groups of exposures. For each group *g* = 1, …, *G*, let *x*_*g*_ denote the design matrix of the exposure variables in the group *g*. The objective function is as follows,

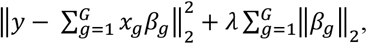

where *β*_*g*_ (*g* = 1, …, *G*) are vector parameters, and ‖·‖_2_ indicates L_2_ norm. In our analysis, we select *λ* value that gives the minimum mean cross-validated error. We implement G-lasso via R package “gglasso” (version 1.5) [37].

#### BKMR

Bayesian kernel machine regression (BKMR) was proposed by Bobb et al. in 2015 [18]. It is a machine learning approach developed to address the statistical challenges in estimating the simultaneous effects from exposure to multiple pollutants. We consider the following model,

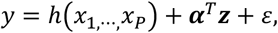

where *h*(·) is unknown function of exposures to be estimated. Covariates ***z*** enter the model linearly, however, if nonlinearity between covariates and outcome are suspected, covariates can also be added into the *h*(·) function to improve overall prediction accuracy and selection. BKMR performs pollutant selection by providing the posterior inclusion probability (PIP, between 0 and 1) for each variable considered in the *h*(·) function. The PIP can be viewed as an importance score, where a higher PIP indicates greater importance of the variable. BKMR does not directly conduct variable selection as the shrinkage method, thus, researchers need to prespecify a threshold value for PIP (e.g., 0.50) to select pollutants with PIPs higher than the threshold. Another aspect to note when using BKMR is that despite BKMR fitting nonlinear functions to the exposures that account for non-additivity, one cannot easily draw conclusions about interaction selection. Therefore, BKMR cannot be evaluated for comparisons of interaction selection accuracy among different methods. In BKMR, one needs to specify the number of iterations of the Markov Chain Monte Carlo (MCMC) sampler, and we run 2000 iterations for all the simulations and 5000 iterations for data analysis. We implement BKMR via R package “bkmr” (version 0.2.0) [38].

#### RF

Random Forest (RF) was proposed by Breiman in 2001 [13]. It is an ensemble learning method for classification that combines randomly generated tree-structured classifiers. It has been successfully applied in a broad range of areas including environmental health. It is more robust to outliers and noise than other methods and it provides variable importance score. For RF, setting a threshold value as in BKMR would be challenging as the importance score describes the accuracy loss if a pollutant is removed. To carry out a pollutant selection function, we propose using k-means clustering [39] to group the pollutant importance into two clusters, and the pollutants in the cluster with higher importance scores are the ones selected. It is important to note that for all other methods except RF, the predictions are based on the selected variables and their model fitting. However, for RF, the variables selected by k-means are meant only as guidelines, as RF is not designed as a variable selection tool. The prediction is based on the model fitting using all the pollutants, not just the selected ones. We implement RF via R package “randomForest” (version 4.6-14) [40].

### Group 2 Methods

This group of approaches aims to perform interaction detection in addition to main effect selection.

#### HierNet

A Lasso for Hierarchical Interactions (HierNet) was proposed by Bien in 2013 [41]. HierNet extends the Lasso for selecting linear main and interaction effects under heredity constraints. The user can choose between strong or weak heredity constraints. With strong heredity, an interaction term is selected into the model only when both of its corresponding main terms are selected, whereas with weak heredity constraint, an interaction term is selected only at least one of its corresponding main terms is selected.

Apart from selecting interactions, HierNet also automatically screens for quadratic terms. However, since quadratic terms are not the primary interest, they are not summarized in this paper. We implement HierNet via R package “hierNet” (version 1.9) [19].

#### HigLasso

Hierarchical Integrative G-Lasso (HigLasso) was proposed by Boss et al. in 2021 [14]. It explores the nonlinear associations between exposures and health outcomes and has been developed as a general shrinkage method that selects nonlinear main and interaction effects of exposures. To specify complex nonlinear relationships, HigLasso adopts a basis expansion approach and it also assumes strong heredity constraint. HigLasso performs variable selection by imposing sparsity on coefficient estimates using G-Lasso penalties. We implement the HigLasso via R package “higlasso” (version 0.9.0) [42].

#### SNIF

Selection of nonlinear interactions by a forward stepwise algorithm (SNIF) was proposed by Narisetty et al. in 2019 [15]. It was motivated by the need to identify chemical mixtures that affect health outcomes and was developed for a general regression model with interaction effects. Like HigLasso, SNIF adopts a basis expansion approach for modeling the nonlinear exposure effects and performs interaction effects selection under the strong heredity constraint. However, SNIF has additional flexibility to retain linear effects of exposures in its selection path, which helps effectively reduce the number of parameters in the model when the linear model fits data well. We implement SNIF via R package “snif” (version 0.5.0) [43].

### Group 3 Methods

This group of approaches aims to optimize the accuracy of predictions for health outcomes while constructing summary risk measures from exposure to chemical mixtures and covariates.

#### ERS

The classic Environmental Risk Score (ERS) was proposed by Park et al. in 2014 and 2017 [23, 24]. It is constructed as a one-dimensional risk score through various predictive models. However, a limitation of this ERS is that it has been restricted to the estimated health effects due to pollutants. The penalization methods such as Lasso, Enet and G-Lasso, can be used to compute the classic ERS as the weighted sum of pollutants or their interactions while controlling for covariates. However, the cutting-edge machine learning algorithms such as BKMR, RF and SNIF are not readily applicable for computing the classic ERS, because they estimate health effects from pollutants and covariates in complex functions without direct separation of pollutant-only effects from other effects. Thus, in this paper we redefine the ERS concept as the prediction for the continuous health outcome or the logit of the probability for the binary outcome. With this updated definition, we can compare ERSs computed through various statistical methods in terms of predictive power, interpretability, and risk stratification. Consider the following model *y* = *g*(*x*_1_, …, *x*_*P*_, *z*_1_, …, *z*_*K*_) + *ε, ε*∼*N*(0, *σ*^2^), we use training data to obtain 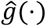, and define ERS as 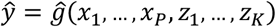 for a collection of pollutants and covariates.

We have selected four methods to construct ERS, namely Enet, BKMR, HierNet, and SNIF, each with distinct strengths in modeling: linear data with easy interpretation (Enet), nonlinear exposure-response relationship (BKMR), linear data with interaction detection (HierNet) and nonlinear data with interaction detection (SNIF). We construct four ERSs, i.e., ERS_Enet_, ERS_BKMR_, ERS_HierNet_ and ERS_SNIF_. To enhance the prediction accuracy of these four ERSs, inspired by the concept of Super Learner (SL [20]), we then construct ERS_SL_ as W_Enet_ERS_Enet_+ W_BKMR_ERS_BKMR_+ W_HierNet_ERS_HierNet_ + W_SNIF_ERS_SNIF_, where the unknown weights W_Enet_, W_BKMR_, W_HierNet_ and W_SNIF_ each range from 0 and 1 and sum to 1. We iteratively solve for these weights using a coordinate descent algorithm. The SL construction is explained below.

#### SL

Super Learner (SL) was proposed by Van der Laan et al. in 2007 [20]. It is a prediction algorithm which aims to find an optimal combination for a collection of candidate learners to minimize the overall risk. On the training dataset with sample size *N*_*train*_, we use ten-fold cross-validation and split the data into 60% for estimation and 40% for validation. We then apply the coordinate descent algorithm with constraint that the four weights are nonnegative and sum to 1. For the *t*th fold (*t* = 1, …,10), let 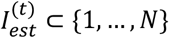 and 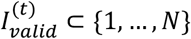 represent the 60% and 40% indices of subjects that are randomly drawn from the training data for estimation and validation respectively, i.e., 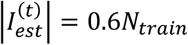 and 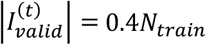, where |·| represents the cardinality of the set. For each learner *j* (*j* = 1, ⋯, 4), we estimate *g* (·) by using the *t*th fold estimation data 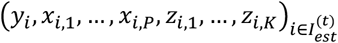 denoted as 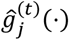. To obtain the weights of the learner, we minimize the loss function on the validation data for continuous outcome, i.e 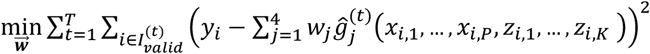, where 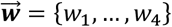. For binary outcome, we use a cross-entropy loss function. Once weights 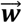 are obtained from training data, on the testing data, we compute 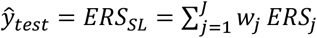, where 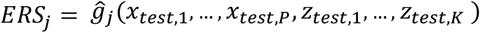 is the ERS independently constructed for each learner *j* (*j* = 1, ⋯, 4) on the testing data.

#### WQS

Weighted quantile sum regression (WQS) was proposed by Carrico et al. in 2015 [21]. It aims to estimate a single dimension disease risk score, called the WQS index, from exposure to mixtures of chemicals. The WQS index is calculated as a weighted sum of individual exposure quantiles. The model is given below,

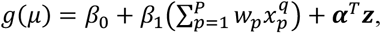

where *g*(·) is the link function in generalized linear model [44] and *μ* is the mean outcome. 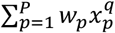 is defined as WQS index, and the weights *w*_*p*_ (*p* = 1, … *P*) are estimated using bootstrapping on training data, which comprises 40% of the total samples by default using R package “gWQS” (version 3.0.4) [45, 46]. The weights *w*_*p*_ are between 0 and 1 and sum to 1. The categorical variable 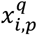 is determined by the quantile of *p*th pollutant exposure for *i*th subject. The quantile transformation enjoys the advantage of standardizing the exposures, and hence the weights describe the relative contribution of each chemical to the joint effect of the health outcome. The remaining 60% of the data is used to test the significance of the coefficient *β*_1_ for the WQS index on the health outcome. The model can also include a set of covariates ***z*** which enter the model linearly and *α*^*T*^ denotes a vector of regression coefficients.

WQS offers a straightforward interpretation by creating a summary index that captures the joint effect of multiple pollutants as well as the relative importance characterized by the magnitude of the weights. However, the validity of directional homogeneity assumption that assumes all the components in the mixture share the same direction of associations with the outcome, should be carefully considered. In addition, transforming continuous pollutant exposures into categorical ones may potentially lead to a loss of information and changes in the correlation structure among pollutants and their true association with the outcome. The package allows users to include interaction terms or quadratic terms of the WQS index to characterize nonlinear association, but these interactions are usually treated as covariates rather than pollutant effects of primary interest.

#### Q-gcomp

Quantile g-computation (Q-gcomp) was proposed by Keil et al. in 2020 [22]. It extends the framework of WQS by relaxing the assumption of directional homogeneity and allowing for positive and negative effects of pollutants. The model without covariates is given by,

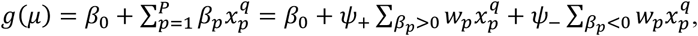

where 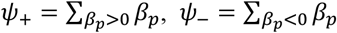, and 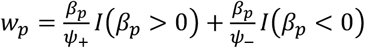. A linear regression model is fit to obtain the coefficients *β*_*p*_ (*p* = 1, … *P*) that determine the estimate of *ψ* = *ψ*___ + *ψ*___ for the summary index and weight *w*_*p*_ for each chemical. The parameter estimation procedure for WQS and Q-gcomp differs in that WQS first estimates the weights *w*_*p*_ on the training data and then estimates *ψ* and its p-value on the validation data based on the estimated weights; while Q-gcomp use all the data to estimate *β*_*p*_ and obtain *ψ*. We implement Q-gcomp via R package “qgcomp” (version 2.8.5) [47].

While both WQS and Q-gcomp provide meaningful summary risk scores from mixtures and rank exposure importance by chemical weights, they do not offer variable selection, potentially limiting their effectiveness in high-dimensional settings. ERS, on the other hand, has two advantages over WQS and Q-gcomp. First, ERS can be constructed using a wide range of statistical prediction approaches, allowing us to incorporate methods with distinct strength to create candidate ERSs that address questions such as variable selection and interaction detection. These candidate ERSs can then be combined to obtain a weighted ERS, referred to as ERS_SL_ to achieve better outcome prediction. Second, since each ERS is constructed using pollutant measurements rather than quantiles, it does not lose any information from the pollutants, leading to more accurate prediction and association detection.

## Simulation Study

### Simulated Data Settings

The simulation studies aim to investigate the associations between chemical mixtures, their interactions and the continuous or binary health outcomes under settings of cross-sectional studies. The 20 pollutants are partitioned into three groups of size seven, six and seven, respectively. For continuous outcomes, we simulate 20 exposures *x*_1_, …, *x*_20_ from a multivariate normal distribution, with mean zero and the marginal variance one. True features are *x*_1_ and *x*_2_; *x*_3_ and *x*_4_; and *x*_5_ from three groups, reflecting that each group includes important toxins. The correlation matrix is specified according to the grouping structure, with within-group correlations and between-group correlations set to 0.6 and 0.1, respectively. Figure 3 shows the heatmap of Pearson correlations among the simulated 20 pollutants. Let *q* = 5 denote the number of true features *x*_1_ to *x*_5_. In settings of LMI and NMI, the 10 pairwise interactions are also true features of outcome *y*. We adopt the specific mean function *g*(·) forms from Boss et al. [48]. A full list of simulation settings and parameters specifications can be found in Table 8.

**Figure 3:**
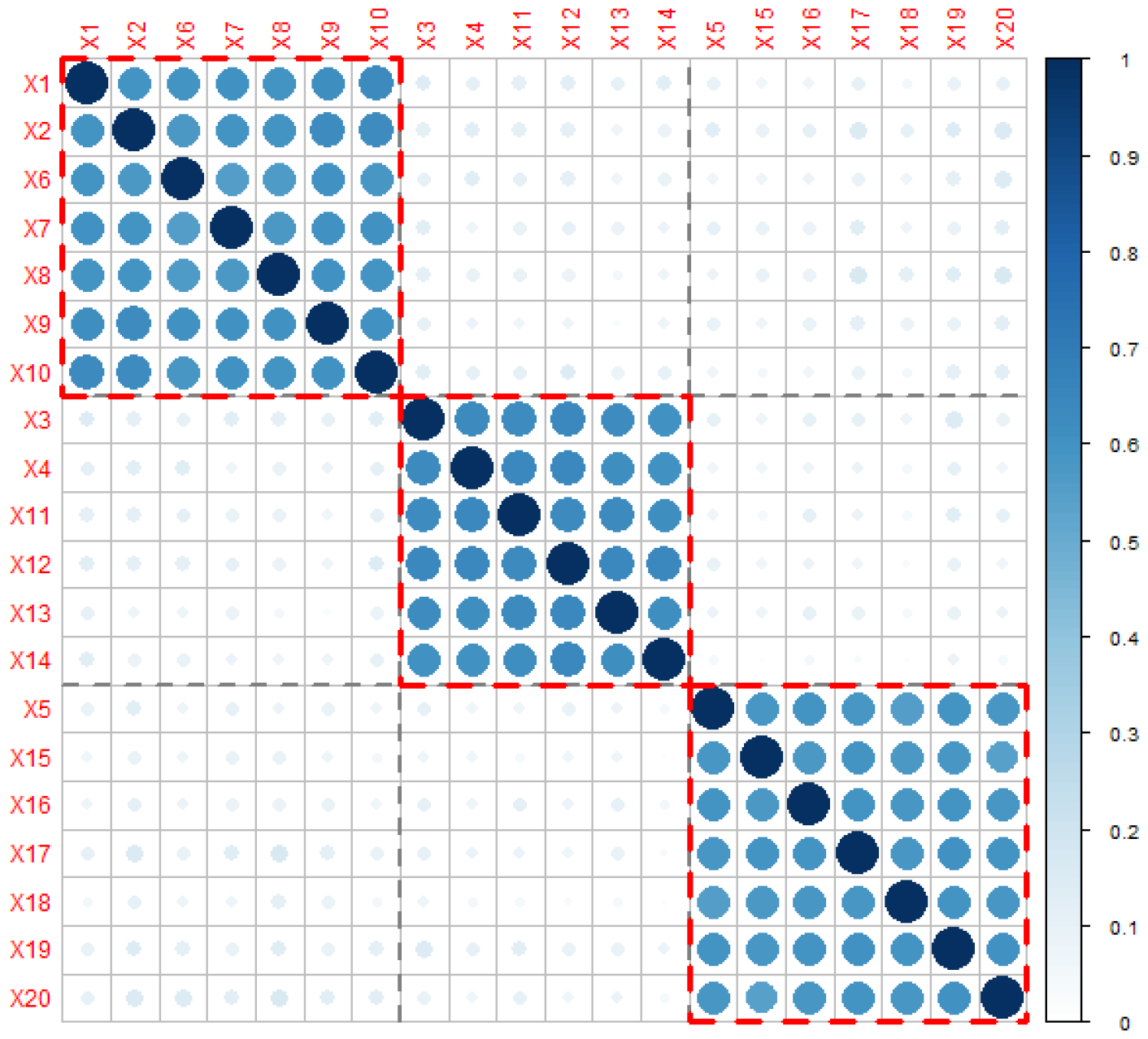
Heat map of Pearson correlations among 20 simulated pollutants with sample size of 1000 and the within-group correlations and between-group correlations are set to 0.6 and 0.1.

#### Evaluation Criteria

Under our analytical framework for analyzing the multipollutant mixture, we utilize standard criteria to evaluate the performance of a collection of statistical methods. For each of the 500 datasets, we randomly spilt 1,000 samples into training and testing datasets, each with 500 samples (i.e., *N*_*train*_= *N*_*test*_ = 500). We evaluate the feature selection and interaction detection on the training dataset and compare the predictive power of three summary scores on the testing dataset. In each dataset under the continuous outcome setting, to assess the feature selection and interaction detection, we consider 20 pollutants and their 190 pairwise interactions, totaling 210 predictors for Lasso, Enet, and G-Lasso (Lasso-MI/Enet-MI/G-Lasso-MI). For comparison purposes, we also fit these three regularization methods with 20 pollutants for main effects only (Lasso-M/Enet-M/G-Lasso-M). We consider 20 pollutants for underlying models of BKMR, RF, HigLasso, HierNet and SNIF, as BKMR and RF do not allow the separation between main and interaction effects, while HigLasso, HierNet and SNIF automatically screen for pairwise interactions for the 20 exposures. For binary outcomes, we have similar settings except that HigLasso and SNIF are no longer available, and BKMR shows unstable simulation results, thus we have to omit these three methods from the analysis of binary outcomes.

To evaluate the accuracy of selecting important pollutants, we use sensitivity, specificity, and false discovery rate (FDR) metrics. These metrics are defined as follows, where *J* = 500 is the number of simulated datasets, and there are 5 true and 15 null effects.

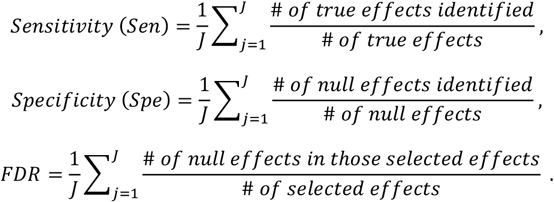

To evaluate the accuracy of interaction detection, we calculate the same three metrics for four settings with interactions: LMI, NMI, LogitI, and NlogitI, where there are 10 true and 180 null interaction effects. In the absence of true interaction effects (settings of LM, NM, Logit, and Nlogit), we utilize the false positive rate (FPR) to assess the proportion of falsely selected interactions out of all 190 interactions. Specifically, FPR is defined as one minus Specificity. The higher the Sensitivity and Specificity values and the lower the FDR and FPR values, the better the feature selection and interaction detection.

For the continuous outcome, to evaluate the prediction performance of three main summary scores (ERS/WQS/Q-gcomp) under various model specifications, we use the testing data. For main effect models, we predict ERS using Enet-M; fit and predict WQS and Q-gcomp models with either pollutants selected by Enet (WQS-M*/Q-gcomp-M*) or all 20 pollutants (WQS-M/Q-gcomp-M). For models considering both main and interactions, we construct ERS using Enet-MI, BKMR, HierNet, SNIF and SL, where the weights of SL are obtained from training data. We also fit and predict WQS and Q-gcomp models with either pollutants and their interactions selected by Enet-MI (WQS-MI*/Q-gcomp-MI*) or all 210 effects (WQS-MI/Q-gcomp-MI).

We evaluate the predictive performance of different methods in three aspects. First, we calculate the Corr and SSE between predicted and observed continuous outcome. Second, to assess the predictive power of summary scores for a binary outcome, we dichotomize the continuous outcome at the 90 percentiles so that the values more than 90 percentiles are 1, calculate AUC to measure the prediction probability of distinguishing between binary outcomes. Third, to evaluate the risk stratification property, we stratify each summary measure on the testing data by the two thresholds of 25 and 75 percentiles of summary measure from the training data. We define the test samples as low (or high) risk group and conduct a logistic regression for these subsets of samples with the dichotomous outcome to obtain an OR of having an extreme outcome between the group with the lowest quartile of the summary measure and the group with the highest quartile of the summary measure. For the binary outcome, in addition to AUC, we also calculate the Brier score defined as the mean of sum of squared errors between the predicted probability and the observed binary outcome. In cases where methods (Enet-M, Enet-MI, BKMR, HierNet and SNIF) fail to select any predictors, we define Corr=0 and OR=1, indicating no predictive power and no risk stratification property from ERS. Figure 4 illustrates the simulation procedure and comparisons among the methods for a continuous outcome, and the procedure for analyzing the binary outcome is similar.

**Figure 4:**
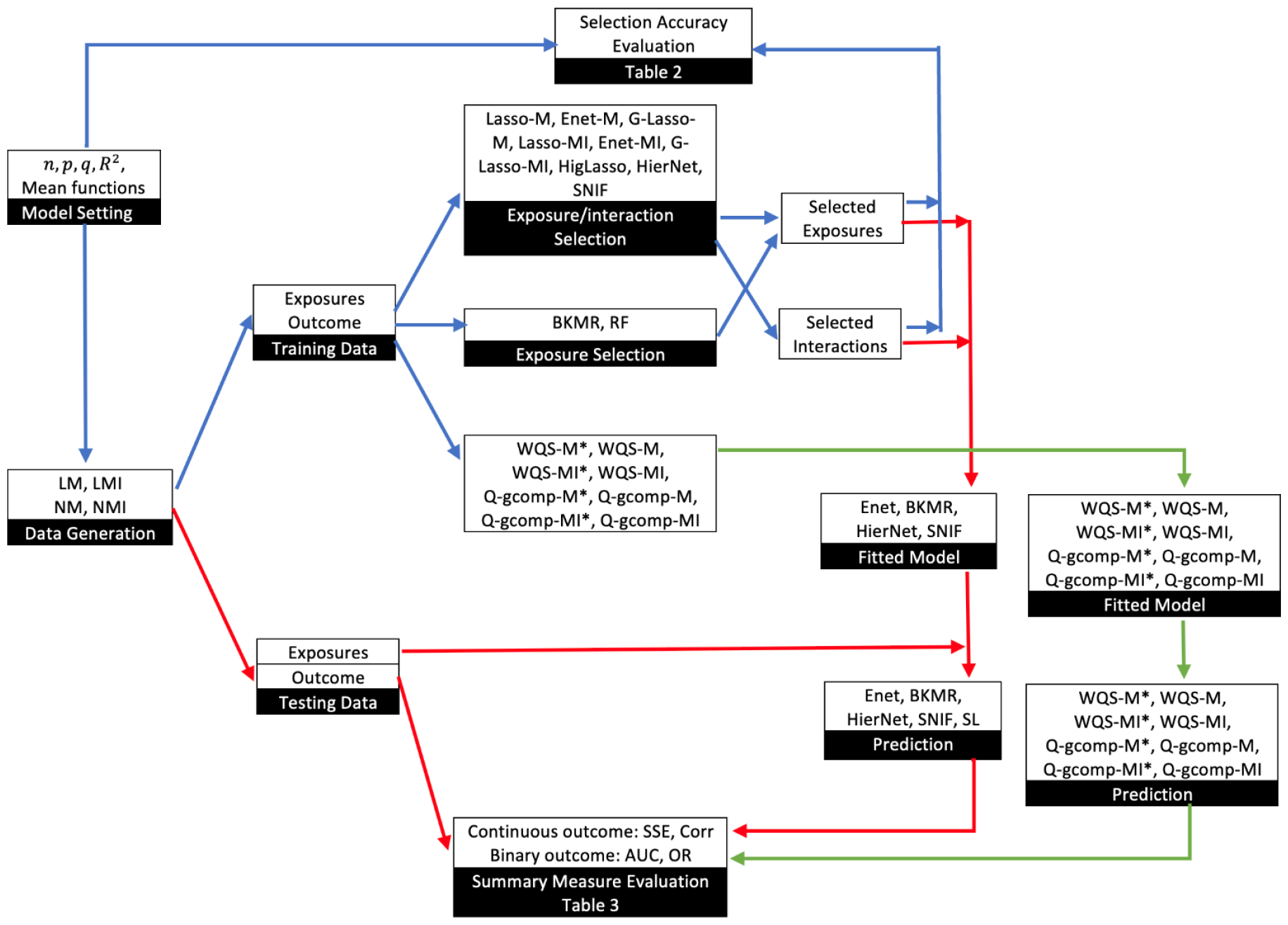
Schematic diagram of the simulation study. *LM* linear main effects, *LMI* linear main effects and interactions, *NM* nonlinear main effects, *NMI* nonlinear main effects and interactions *Lasso-M* lasso for main effects, *Enet-M* elastic net for main effects, *G-Lasso-M* group lasso for main effects, *Lasso-MI* lasso for main effects and interactions, *Enet-MI* elastic net for main effects and interactions, *G-Lasso-MI* group lasso for main effects and interactions, *BKMR* Bayesian kernel machine regression, *RF* random forest, *HigLasso* hierarchical integrative group lasso, *HierNet* lasso for hierarchical interactions, *SNIF* selection of nonlinear interactions by a forward stepwise algorithm, *WQS-M\** weighted quantile sum regression (WQS) for selected main effects by Enet-M, *WQS-M* WQS for main effects, *WQS-MI\** WQS for selected main effects and interactions by Enet-MI, *WQS-MI* WQS for main effects and interactions, *Q-gcomp-M\** quantile g-computation (Q-gcomp) for selected main effects by Enet-M, *Q-gcomp-M* Q-gcomp for main effects, *Q-gcomp-MI\** Q-gcomp for selected main effects and interactions by Enet-MI, *Q-gcomp-MI* Q-gcomp for main effects and interactions

### PROTECT Data Application

#### Data description

We apply the proposed framework to a data analysis from PROTECT study, which aim to determine the impacts of exposure to mixtures from four chemical classes (metals, phthalates, phenols and PAHs) during pregnancy on adverse outcomes of birth weight and preterm birth. Women were eligible to participate if they were between the ages of 18 and 40 years, had their first clinic visit before their 20^th^ week of pregnancy, did not use *in vitro* fertilization to become pregnant, did not use oral contraceptives within three months of becoming pregnant, and had no known preexisting medical or obstetric conditions. The PROTECT study was approved by the research and ethics committees of the University of Michigan School of Public Health, University of Puerto Rico, Northeastern University, and all participating hospitals and clinics. All participants provided full informed consent prior to participation.

#### Exposures, covariates, and outcomes

We start with a dataset that included 61 pollutants, including 19 phthalates, 12 phenols, 8 PAHs and 22 metals, collected from urine samples of 1,747 women during gestation across three visits. We then create reduced datasets including only individuals with complete data on each outcome (birth weight, preterm birth status) and covariates. This result in sample sizes of 1,348 for birth weight (kg) and 1,379 for preterm birth (yes/no). To analyze birth weight as a binary outcome, we dichotomize it at 2.5 kg for low birth weight and at 4.0 kg for high birth weight (formally termed as fetal macrosomia). Among our sample, we have 93 out of 1,348 children with low birth weight (6.90%), 55 out of 1,348 children with high birth weight (4.08%). Preterm birth is defined as gestational age less than 37 weeks at delivery. We have 126 out of 1,379 children born preterm (9.14%). We adjust for covariates of infant sex, education (high school or less, some college, college or above) and maternal age at recruitment (years) for birth weight. We impute exposure concentrations (ng/dL) measured below limit of detection (LOD) with 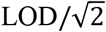, and correct them by urine specific gravity (SG) using the equation 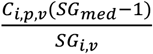, where *SG*_*med*_ is the median urine specific gravity in this dataset (1.019); *SG*_*i,v*_ is the individual *i* urine specific gravity at visit *v*; and *C*_*i,v,p*_ is the *p*-th pollutant concentration for individual *i* at visit *v*. Due to the right-skewed distributions of SG-adjusted concentrations, we apply the logarithmic transformation with base 10 on the concentrations.

After evaluating the percentage of samples measured above LOD for each pollutant, we eliminate 14 pollutants from our analysis, due to their measurements above LOD less than 70% of the samples at any visit. Furthermore, we exclude eight additional pollutants due to missingness in more than 20% of samples after taking the mean across three visits in our dataset. We impute missing values for all remaining chemicals (4.23-15.50% missing) via R package “missForest” (version 1.4) [49, 50] based on single imputation. After these preprocessing steps, our final dataset consists of 39 chemical exposures (14 metals, 7 PAHs, 11 phthalates and 7 phenols), four covariates (three covariates for preterm), and 903 pairwise interactions among all chemicals and covariates.

#### Statistical analysis

For birth weight, we randomly split the 1,348 samples into 674 samples each for training and testing data. For feature selection, we fit Enet with underlying models considering first 39 main effects (Enet-M) adjusting for four covariates. Next, we fit 43 main effects along with their pairwise 903 interactions (Enet-MI). For BKMR, we fit 39 chemicals in the nonlinear function while adjusting for four covariates linearly, with PIP’s cutoff of 0.80. For HierNet and SNIF, we fit the models with 43 main effects, where the HierNet and SNIF screens for main and 903 interaction effects by default. We compare three main effect models that are adjusted for covariates: ERS obtained from Enet (ERS_Enet-M_), WQS (WQS-M) and Q-gcomp (Q-gcomp-M). We don’t fit WQS and Q-gcomp with the main effects selected from Enet-M as in our simulations, because Enet fails to select any chemicals in 22 of the 100 training datasets. For main and interaction effect models, we compare five ERSs (Enet-MI, BKMR, HierNet, SNIF and SL) on the testing data using following metrics. First, we calculate Corr and SSE between observed birth weight and ERSs. Second, we use low birth weight (Yes=1/No=0) as the binary outcome and calculate AUC to evaluate the prediction power of ERSs. Third, we categorize ERSs from the testing data using Q_1_ obtained from training data and define the subjects with ERS below Q_1_ as high risk group, and compute the OR of low birth weight between the high risk group and the rest of the samples to evaluate the risk discrimination ability of each ERS. For high birth weight, we calculate the OR of high birth weight for subjects with ERS more than Q_3_ versus the rest of the subjects. Due to the uncertainties with each random split of the samples, we repeat the entire model fitting and validation procedure 100 times and report the pollutants and interactions that are consistently selected by each method for at least 30% of the 100 times. For preterm birth (Yes=1/No=0), the analysis is similar and for main and interaction models, we compare five ERSs (Lasso-MI, Enet-MI, RF, HierNet and SL), report brier scores and define ERS less than Q_1_ as low risk group and higher than Q_3_ as high-risk group to reflect the higher the ERS, the higher the probability of a preterm birth.

## Data Availability

We used the existing datasets collected in PROTECT cohort study(P42ES017198). This is de-identified data. For researchers interested in using this data, they will need to contact the Principal Investigators of PROTECT study for data access.

## Acknowledgements

This research work was supported by NIH-funded longitudinal ongoing cohort study “Puerto Rico PROTECT birth cohort” (P42ES017198).

## Notes

### Competing Interest Statement

The authors have declared no competing interest.

### Author Declarations

The PROTECT study was approved by the research and ethics committees of the University of Michigan School of Public Health, University of Puerto Rico, Northeastern University, and all participating hospitals and clinics.

