## supplementary info for "Statistical methods for chemical mixtures: a roadmap for practitioners"

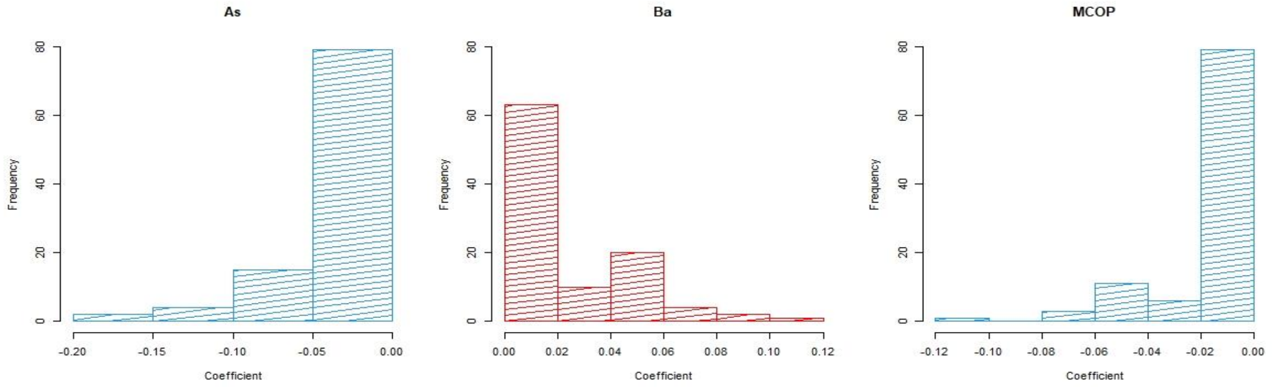


**Figure S1: Distributions of coefficient estimates for chemicals Ba, As and MCOP by Enet-M**

**Table S1: Selection accuracy for main and interaction identification among eight methods, where outcome is generated from LM, NM, LMI, and NMI. Means of Sen, Spe, FDR and FPR are obtained from 100 data replications with $N_{train} = 500$, $p = 20$, $q = 5$, and $R^2 = 0.1$**

| Data | Type | Metric | Lasso-M | Enet-M | G-Lasso-M | Lasso-MI | Enet-MI | G-Lasso-MI | BKMR | RF | HigLasso | HierNet | SNIF |
|---|---|---|---|---|---|---|---|---|---|---|---|---|---|
| LM | Main/Marginal | Sen | 0.856 | 0.866 | 1.000 | 0.788 | 0.810 | 1.000 | 1.000 | 0.550 | 0.510 | 0.866 | 0.476 |
| | | Spe | 0.661 | 0.642 | 0.369 | 0.790 | 0.759 | 0.000 | 0.001 | 0.804 | 0.861 | 0.587 | 0.955 |
| | | FDR | 0.521 | 0.534 | 0.602 | 0.421 | 0.450 | 0.750 | 0.750 | 0.443 | 0.330 | 0.564 | 0.204 |
| | Interaction | FPR | -- | -- | -- | 0.042 | 0.051 | 1.000 | -- | -- | 0.004 | 0.057 | 0.000 |
| LMI | Main/Marginal | Sen | 0.454 | 0.528 | 0.920 | 0.432 | 0.468 | 1.000 | 1.000 | 0.508 | 0.342 | 0.900 | 0.296 |
| | | Spe | 0.771 | 0.724 | 0.438 | 0.820 | 0.803 | 0.000 | 0.004 | 0.767 | 0.893 | 0.423 | 0.945 |
| | | FDR | 0.519 | 0.583 | 0.569 | 0.532 | 0.538 | 0.750 | 0.749 | 0.518 | 0.396 | 0.626 | 0.307 |
| | Interaction | Sen | -- | -- | -- | 0.377 | 0.407 | 1.000 | -- | -- | 0.077 | 0.297 | 0.030 |
| | | Spe | -- | -- | -- | 0.918 | 0.906 | 0.000 | -- | -- | 0.991 | 0.938 | 1.000 |
| | | FDR | -- | -- | -- | 0.781 | 0.789 | 0.947 | -- | -- | 0.498 | 0.756 | 0.070 |
| NM | Main/Marginal | Sen | 0.556 | 0.576 | 1.000 | 0.474 | 0.500 | 1.000 | 0.846 | 0.446 | 0.556 | 0.810 | 0.480 |
| | | Spe | 0.713 | 0.672 | 0.331 | 0.827 | 0.795 | 0.000 | 0.264 | 0.831 | 0.899 | 0.562 | 0.965 |
| | | FDR | 0.560 | 0.598 | 0.621 | 0.475 | 0.521 | 0.750 | 0.633 | 0.408 | 0.220 | 0.594 | 0.142 |
| | Interaction | FPR | -- | -- | -- | 0.074 | 0.085 | 1.000 | -- | -- | 0.007 | 0.051 | 0.000 |
| NMI | Main/Marginal | Sen | 0.562 | 0.594 | 0.990 | 0.496 | 0.510 | 1.000 | 0.924 | 0.474 | 0.512 | 0.840 | 0.454 |
| | | Spe | 0.717 | 0.682 | 0.345 | 0.821 | 0.791 | 0.000 | 0.128 | 0.829 | 0.909 | 0.512 | 0.961 |
| | | FDR | 0.558 | 0.586 | 0.607 | 0.483 | 0.521 | 0.750 | 0.703 | 0.430 | 0.237 | 0.614 | 0.167 |
| | Interaction | Sen | -- | -- | -- | 0.193 | 0.207 | 1.000 | -- | -- | 0.080 | 0.151 | 0.004 |
| | | Spe | -- | -- | -- | 0.924 | 0.919 | 0.000 | -- | -- | 0.997 | 0.945 | 1.000 |
| | | FDR | -- | -- | -- | 0.860 | 0.865 | 0.947 | -- | -- | 0.184 | 0.853 | 0.010 |

*LM* linear main effects, *LMI* linear main effects and interactions, *NM* nonlinear main effects, *NMI* nonlinear main effects and interactions
*Sen* sensitivity, *Spe* specificity, *FDR* false discovery rate, *FPR* false positive rate
*Lasso-M* lasso for main effects, *Enet-M* elastic net for main effects, *G-Lasso-M* group lasso for main effects, *Lasso-MI* lasso for main effects and interactions, *Enet-MI* elastic net for main effects and interactions, *G-Lasso-MI* group lasso for main effects and interactions, *BKMR* Bayesian kernel machine regression, *RF* random forest, *HigLasso* hierarchical integrative group lasso, *HierNet* lasso for hierarchical interactions, *SNIF* selection of nonlinear interactions by a forward stepwise algorithm

**Table S2: Risk prediction performance by different statistical methods, when data are generated from LM, NM, LMI, and NMI. Means of Corr, SSE, AUC, and median of OR are obtained from 100 data replications for $N_{test} = 500, p = 20, q = 5$, and $R^2 = 0.1$**

| Data | Metric | ERS Enet-M | WQS -M* | WQS -M | Q-gcomp -M* | Q-gcomp -M | ERS Enet-MI | ERS BKMR | ERS HierNet | ERS SNIF | ERS SL | WQS -MI* | WQS -MI | Q-gcomp -MI* | Q-gcomp -MI |
|---|---|---|---|---|---|---|---|---|---|---|---|---|---|---|---|
| | | **Continuous Outcome and Continuous ERS/WQS/Q-gcomp** | | | | | | | | | | | | | |
| LM | Corr | 0.28 | 0.27 | 0.27 | 0.26 | 0.25 | 0.26 | 0.12 | 0.26 | 0.25 | 0.26 | 0.24 | 0.20 | 0.23 | 0.10 |
| | SSE | 83.0 | 83.6 | 83.7 | 84.3 | 85.4 | 84.1 | 91.6 | 83.9 | 86.0 | 83.9 | 85.2 | 86.8 | 88.0 | 147.9 |
| LMI | Corr | 0.11 | 0.12 | 0.12 | 0.11 | 0.09 | 0.24 | 0.08 | 0.21 | 0.14 | 0.24 | 0.22 | 0.23 | 0.20 | 0.09 |
| | SSE | 370.7 | 371.4 | 370.1 | 374.8 | 381.1 | 353.0 | 387.3 | 358.9 | 375.5 | 353.6 | 359.9 | 356.2 | 377.6 | 618.3 |
| NM | Corr | 0.20 | 0.18 | 0.18 | 0.17 | 0.16 | 0.22 | 0.18 | 0.25 | 0.23 | 0.25 | 0.19 | 0.18 | 0.17 | 0.08 |
| | SSE | 157.2 | 158.7 | 158.7 | 160.2 | 162.8 | 155.7 | 163.7 | 153.5 | 161.1 | 153.7 | 159.6 | 158.8 | 167.2 | 274.5 |
| NMI | Corr | 0.19 | 0.17 | 0.17 | 0.16 | 0.15 | 0.22 | 0.14 | 0.24 | 0.20 | 0.24 | 0.20 | 0.21 | 0.17 | 0.07 |
| | SSE | 240.5 | 242.6 | 242.2 | 244.8 | 248.3 | 236.5 | 256.5 | 234.8 | 247.6 | 234.7 | 241.7 | 239.2 | 253.3 | 416.6 |
| | | **Dichotomous Outcome and Continuous ERS/ WQS/Q-gcomp** | | | | | | | | | | | | | |
| LM | AUC | 0.65 | 0.65 | 0.64 | 0.64 | 0.64 | 0.64 | 0.56 | 0.64 | 0.63 | 0.64 | 0.63 | 0.61 | 0.63 | 0.56 |
| LMI | AUC | 0.59 | 0.59 | 0.59 | 0.59 | 0.57 | 0.64 | 0.56 | 0.62 | 0.58 | 0.64 | 0.63 | 0.63 | 0.62 | 0.57 |
| NM | AUC | 0.62 | 0.61 | 0.61 | 0.61 | 0.60 | 0.63 | 0.61 | 0.65 | 0.63 | 0.64 | 0.61 | 0.61 | 0.60 | 0.55 |
| NMI | AUC | 0.62 | 0.61 | 0.61 | 0.61 | 0.60 | 0.63 | 0.59 | 0.64 | 0.62 | 0.64 | 0.62 | 0.62 | 0.61 | 0.56 |
| | | **Dichotomous Outcome and Categorical ERS/ WQS/Q-gcomp** | | | | | | | | | | | | | |
| LM | OR | 4.2 | 4.3 | 3.7 | 4.3 | 4.0 | 4.0 | 2.2 | 3.8 | 3.4 | 3.9 | 3.5 | 2.7 | 3.1 | 1.7 |
| LMI | OR | 2.0 | 2.1 | 2.0 | 1.9 | 1.9 | 3.1 | 1.5 | 2.6 | 2.2 | 3.7 | 3.1 | 3.0 | 2.6 | 1.9 |
| NM | OR | 2.8 | 2.7 | 2.7 | 2.7 | 2.5 | 3.1 | 2.8 | 3.5 | 3.2 | 3.5 | 2.9 | 2.5 | 2.5 | 1.6 |
| NMI | OR | 2.7 | 2.6 | 2.6 | 2.6 | 2.5 | 3.0 | 2.4 | 3.1 | 2.8 | 3.3 | 2.7 | 3.0 | 2.7 | 1.6 |

*LM* linear main effects, *LMI* linear main effects and interactions, *NM* nonlinear main effects, *NMI* nonlinear main effects and interactions
*Corr* correlation, *SSE* sum of squared error, *AUC* area under the receiver operating characteristic curve, *OR* odds ratio
*Enet-M* elastic net for main effects, *WQS-M** weighted quantile sum regression (WQS) for selected main effects by Enet-M, *WQS-M* WQS for main effects, *Q-gcomp-M** quantile g-computation (Q-gcomp) for selected main effects by Enet-M, *Q-gcomp-M* Q-gcomp for main effects, *Enet-MI* elastic net for main effects and interactions, *BKMR* Bayesian kernel machine regression, *HierNet* lasso for hierarchical interactions, *SNIF* selection of nonlinear interactions by a forward stepwise algorithm, *SL* super learner, *WQS-MI** WQS for selected main effects and interactions by Enet-MI, *WQS-MI* WQS for main effects and interactions, *Q-gcomp-Mi** Q-gcomp for selected main effects and interactions by Enet-MI, *Q-gcomp-MI* Q-gcomp for main effects and interactions

**Table S3: Selection accuracy for main and interaction identification among five methods, where outcome is generated from Logit, LogitI, NLogitI, and NLogitI. Means of Sen, Spe, FDR and FPR are obtained from 100 data replications with $N_{train} = 500, p = 20, q = 5$ and $R^2 = 0.1$**

| Data | Type | Metric | Lasso-M | Enet-M | G-Lasso-M | Lasso-MI | Enet-MI | G-Lasso-MI | RF | HierNet |
|---|---|---|---|---|---|---|---|---|---|---|
| Logit | Main/Marginal | Sen | 0.858 | 0.890 | 0.830 | 0.772 | 0.838 | 0.960 | 0.532 | 0.548 |
| | | Spe | 0.676 | 0.607 | 0.447 | 0.829 | 0.757 | 0.040 | 0.656 | 0.752 |
| | | FDR | 0.502 | 0.552 | 0.498 | 0.363 | 0.440 | 0.720 | 0.626 | 0.381 |
| | Interaction | FPR | -- | -- | -- | 0.043 | 0.060 | 0.960 | -- | 0.034 |
| LogitI | Main/Marginal | Sen | 0.796 | 0.834 | 0.860 | 0.646 | 0.734 | 0.930 | 0.462 | 0.658 |
| | | Spe | 0.701 | 0.641 | 0.330 | 0.829 | 0.777 | 0.070 | 0.657 | 0.608 |
| | | FDR | 0.505 | 0.545 | 0.585 | 0.407 | 0.444 | 0.698 | 0.665 | 0.519 |
| | Interaction | Sen | -- | -- | -- | 0.140 | 0.177 | 0.930 | -- | 0.136 |
| | | Spe | -- | -- | -- | 0.948 | 0.928 | 0.070 | -- | 0.950 |
| | | FDR | -- | -- | -- | 0.841 | 0.859 | 0.881 | -- | 0.606 |
| Nlogit | Main/Marginal | Sen | 0.602 | 0.636 | 0.830 | 0.494 | 0.548 | 0.940 | 0.408 | 0.654 |
| | | Spe | 0.751 | 0.685 | 0.323 | 0.875 | 0.801 | 0.060 | 0.733 | 0.669 |
| | | FDR | 0.508 | 0.578 | 0.617 | 0.383 | 0.494 | 0.705 | 0.569 | 0.475 |
| | Interaction | FPR | -- | -- | -- | 0.051 | 0.074 | 0.940 | -- | 0.044 |
| NlogitI | Main/Marginal | Sen | 0.600 | 0.658 | 0.810 | 0.514 | 0.562 | 0.920 | 0.402 | 0.670 |
| | | Spe | 0.748 | 0.675 | 0.316 | 0.871 | 0.794 | 0.080 | 0.699 | 0.603 |
| | | FDR | 0.520 | 0.574 | 0.579 | 0.380 | 0.497 | 0.690 | 0.622 | 0.521 |
| | Interaction | Sen | -- | -- | -- | 0.118 | 0.157 | 0.920 | -- | 0.121 |
| | | Spe | -- | -- | -- | 0.948 | 0.926 | 0.080 | -- | 0.950 |
| | | FDR | -- | -- | -- | 0.877 | 0.877 | 0.872 | -- | 0.657 |

*Logit* logit-link linear main effects, *LogitI* logit-link linear main effects and interactions, *Nlogit* logit-link nonlinear main effects, *NlogitI* logit-link nonlinear main effects and interactions
*Sen* sensitivity, *Spe* specificity, *FDR* false discovery rate, *FPR* false positive rate
*Lasso-M* lasso for main effects, *Enet-M* elastic net for main effects, *G-Lasso-M* group lasso for main effects, *Lasso-MI* lasso for main effects and interactions, *Enet-MI* elastic net for main effects and interactions, *G-Lasso-MI* group lasso for main effects and interactions, *RF* random forest, *HierNet* lasso for hierarchical interactions
Mean prevalence of outcome over 100 replicates equals to 15.2%, 15.1%, 16.4% and 15.4% for Logit, LogitI, Nlogit and NlogitI, respectively

**Table S4: Risk prediction performance by different statistical methods, when data are generated from Logit, LogitI, Nlogit, and NLogitI. Means of AUC, Brier, and median of OR are obtained from 100 data replications for $N_{test} = 500, p = 20, q = 5$ and $R^2 = 0.1$**

| Data | Metric | ERS Enet-M | WQS -M* | WQS -M | Q-gcomp -M* | Q-gcomp -M | ERS Lasso-MI | ERS Enet-MI | ERS RF | ERS HierNet | ERS SL | WQS -MI* | WQS -MI | Q-gcomp -MI* | Q-gcomp -MI |
|---|---|---|---|---|---|---|---|---|---|---|---|---|---|---|---|
| | | **Dichotomous Outcome and Continuous ERS WQS/Q-gcomp** | | | | | | | | | | | | | |
| Logit | AUC | 0.720 | 0.714 | 0.715 | 0.707 | 0.696 | 0.705 | 0.707 | 0.681 | 0.681 | 0.708 | 0.683 | 0.656 | 0.676 | 0.548 |
| | Brier | 0.118 | 0.119 | 0.120 | 0.121 | 0.123 | 0.120 | 0.120 | 0.123 | 0.124 | 0.123 | 0.122 | 0.123 | 0.128 | 0.314 |
| LogitI | AUC | 0.686 | 0.679 | 0.680 | 0.675 | 0.661 | 0.665 | 0.667 | 0.649 | 0.659 | 0.669 | 0.651 | 0.638 | 0.642 | 0.543 |
| | Brier | 0.120 | 0.121 | 0.121 | 0.122 | 0.124 | 0.121 | 0.121 | 0.123 | 0.123 | 0.122 | 0.122 | 0.123 | 0.128 | 0.316 |
| Nlogit | AUC | 0.709 | 0.694 | 0.688 | 0.690 | 0.673 | 0.701 | 0.699 | 0.683 | 0.698 | 0.704 | 0.669 | 0.655 | 0.662 | 0.541 |
| | Brier | 0.125 | 0.128 | 0.128 | 0.129 | 0.132 | 0.126 | 0.127 | 0.129 | 0.128 | 0.127 | 0.130 | 0.131 | 0.136 | 0.325 |
| NlogitI | AUC | 0.704 | 0.688 | 0.689 | 0.684 | 0.669 | 0.696 | 0.695 | 0.675 | 0.690 | 0.698 | 0.664 | 0.650 | 0.657 | 0.542 |
| | Brier | 0.120 | 0.122 | 0.122 | 0.123 | 0.125 | 0.120 | 0.120 | 0.123 | 0.122 | 0.121 | 0.123 | 0.124 | 0.129 | 0.317 |
| | | **Dichotomous Outcome and Categorical ERS WQS/Q-gcomp** | | | | | | | | | | | | | |
| Logit | OR | 8.7 | 8.1 | 8.0 | 7.6 | 6.4 | 7.6 | 7.5 | 4.8 | 5.8 | 8.1 | 5.1 | 3.8 | 5.4 | 1.5 |
| LogitI | OR | 5.3 | 4.9 | 4.9 | 4.8 | 4.4 | 4.2 | 4.4 | 2.5 | 4.2 | 4.3 | 3.5 | 3.0 | 3.7 | 1.4 |
| Nlogit | OR | 7.4 | 6.1 | 5.8 | 6.5 | 5.2 | 6.9 | 6.6 | 4.9 | 6.1 | 7.2 | 4.7 | 3.9 | 4.4 | 1.5 |
| NlogitI | OR | 7.0 | 5.5 | 5.6 | 5.9 | 4.9 | 6.3 | 6.1 | 4.2 | 5.5 | 6.2 | 4.4 | 3.7 | 4.1 | 1.4 |

*Logit* logit-link linear main effects, *LogitI* logit-link linear main effects and interactions, *Nlogit* logit-link nonlinear main effects, *NlogitI* logit-link nonlinear main effects and interactions

*AUC* area under the receiver operating characteristic curve, *Brier* Brier score, *OR* odds ratio

*Enet-M* elastic net for main effects, *WQS-M** weighted quantile sum regression (WQS) for selected main effects by Enet-M, *WQS-M* WQS for main effects, *Q-gcomp-M** quantile g-computation (Q-gcomp) for selected main effects by Enet-M, *Q-gcomp-M* Q-gcomp for main effects, *Lasso-MI* lasso for main effects and interactions, *Enet-MI* elastic net for main effects and interactions, *RF* random forest, *HierNet* lasso for hierarchical interactions, *SL* super learner, *WQS-MI** WQS for selected main effects and interactions by Enet-MI, *WQS-MI* WQS for main effects and interactions, *Q-gcomp-Mi** Q-gcomp for selected main effects and interactions by Enet-MI, *Q-gcomp-MI* Q-gcomp for main effects and interactions

Mean prevalence of outcome over 100 replicates equals to 15.2%, 15.1%, 16.4% and 15.4% for Logit, LogitI, Nlogit and NlogitI, respectively

**Table S5 Selection accuracy for main and interaction identification among eight methods, where outcome is generated from LM, NM, LMI, and NMI. Means of Sen, Spe, FDR and FPR are obtained from 100 data replications with $N_{train} = 1000$, $p = 40$, $q = 5$, and $R^2 = 0.2$**

| Data | Type | Metric | Lasso-M | Enet-M | Lasso-MI | Enet-MI | BKMR | RF | HierNet | SNIF |
|---|---|---|---|---|---|---|---|---|---|---|
| LM | Main/Marginal | Sen | 0.998 | 0.998 | 0.994 | 0.996 | 0.998 | 0.698 | 0.998 | 0.906 |
| | | Spe | 0.720 | 0.696 | 0.834 | 0.799 | 0.011 | 0.962 | 0.706 | 0.985 |
| | | FDR | 0.629 | 0.656 | 0.520 | 0.567 | 0.874 | 0.191 | 0.645 | 0.097 |
| | Interaction | FPR | -- | -- | 0.015 | 0.017 | -- | -- | 0.022 | 0.000 |
| LMI | Main/Marginal | Sen | 0.754 | 0.776 | 0.744 | 0.752 | 0.994 | 0.664 | 1.000 | 0.490 |
| | | Spe | 0.760 | 0.752 | 0.841 | 0.828 | 0.020 | 0.916 | 0.225 | 0.963 |
| | | FDR | 0.663 | 0.668 | 0.581 | 0.599 | 0.872 | 0.333 | 0.842 | 0.281 |
| | Interaction | Sen | -- | -- | 0.724 | 0.758 | -- | -- | 0.556 | 0.157 |
| | | Spe | -- | -- | 0.961 | 0.956 | -- | -- | 0.955 | 1.000 |
| | | FDR | -- | -- | 0.793 | 0.804 | -- | -- | 0.859 | 0.172 |
| NM | Main/Marginal | Sen | 0.608 | 0.610 | 0.540 | 0.554 | 0.840 | 0.420 | 0.928 | 0.676 |
| | | Spe | 0.781 | 0.763 | 0.893 | 0.870 | 0.433 | 0.994 | 0.620 | 0.987 |
| | | FDR | 0.653 | 0.685 | 0.539 | 0.583 | 0.660 | 0.042 | 0.725 | 0.096 |
| | Interaction | FPR | -- | -- | 0.035 | 0.039 | -- | -- | 0.027 | 0.000 |
| NMI | Main/Marginal | Sen | 0.654 | 0.672 | 0.596 | 0.608 | 0.932 | 0.464 | 0.972 | 0.710 |
| | | Spe | 0.773 | 0.757 | 0.867 | 0.848 | 0.268 | 0.990 | 0.571 | 0.983 |
| | | FDR | 0.662 | 0.677 | 0.576 | 0.611 | 0.755 | 0.076 | 0.742 | 0.111 |
| | Interaction | Sen | -- | -- | 0.290 | 0.312 | -- | -- | 0.235 | 0.015 |
| | | Spe | -- | -- | 0.962 | 0.958 | -- | -- | 0.972 | 1.000 |
| | | FDR | -- | -- | 0.903 | 0.907 | -- | -- | 0.895 | 0.010 |

*LM* linear main effects, *LMI* linear main effects and interactions, *NM* nonlinear main effects, *NMI* nonlinear main effects and interactions
*Sen* sensitivity, *Spe* specificity, *FDR* false discovery rate, *FPR* false positive rate
*Lasso-M* lasso for main effects, *Enet-M* elastic net for main effects, *Lasso-MI* lasso for main effects and interactions, *Enet-MI* elastic net for main effects and interactions, *BKMR* Bayesian kernel machine regression, *RF* random forest, *HierNet* lasso for hierarchical interactions, *SNIF* selection of nonlinear interactions by a forward stepwise algorithm

**Table S6: Risk prediction performance by different statistical methods, when data are generated from LM, NM, LMI, and NMI. Means of Corr, SSE, AUC, and median of OR are obtained from 100 data replications for $N_{test} = 1000$, $p = 40$, $q = 5$, and $R^2 = 0.2$**

| Data | Metric | ERS Enet-M | WQS -M* | WQS -M | Q-gcomp -M* | Q-gcomp -M | ERS Enet-MI | ERS BKMR | ERS HierNet | ERS SNIF | ERS SL | WQS -MI* | WQS -MI | Q-gcomp -MI* | Q-gcomp -MI |
|---|---|---|---|---|---|---|---|---|---|---|---|---|---|---|---|
| | | **Continuous Outcome and Continuous ERS/WQS/Q-gcomp** | | | | | | | | | | | | | |
| LM | Corr | 0.43 | 0.41 | 0.41 | 0.41 | 0.39 | 0.43 | 0.30 | 0.42 | 0.43 | 0.43 | 0.40 | 0.34 | 0.38 | 0.09 |
| | SSE | 36.4 | 37.1 | 37.2 | 37.4 | 38.1 | 36.7 | 41.2 | 36.7 | 36.5 | 36.4 | 37.8 | 39.4 | 38.4 | 244.1 |
| LMI | Corr | 0.19 | 0.18 | 0.19 | 0.17 | 0.15 | 0.40 | 0.27 | 0.38 | 0.32 | 0.40 | 0.35 | 0.34 | 0.33 | 0.08 |
| | SSE | 175.3 | 176.1 | 175.7 | 177.5 | 181.1 | 152.6 | 172.4 | 155.9 | 173.4 | 152.8 | 160.1 | 160.6 | 167.1 | 1037.2 |
| NM | Corr | 0.32 | 0.29 | 0.28 | 0.28 | 0.25 | 0.39 | 0.36 | 0.41 | 0.40 | 0.42 | 0.32 | 0.30 | 0.30 | 0.06 |
| | SSE | 73.0 | 74.3 | 74.7 | 75.0 | 76.7 | 69.2 | 71.1 | 67.2 | 75.3 | 67.2 | 73.2 | 74.0 | 75.7 | 474.5 |
| NMI | Corr | 0.30 | 0.28 | 0.28 | 0.27 | 0.25 | 0.39 | 0.34 | 0.40 | 0.38 | 0.40 | 0.33 | 0.31 | 0.30 | 0.06 |
| | SSE | 110.4 | 112.3 | 112.5 | 113.1 | 115.6 | 103.9 | 108.0 | 102.1 | 112.6 | 101.9 | 109.0 | 109.8 | 113.5 | 710.9 |
| | | **Dichotomous Outcome and Continuous ERS/ WQS/Q-gcomp** | | | | | | | | | | | | | |
| LM | AUC | 0.74 | 0.73 | 0.73 | 0.73 | 0.72 | 0.74 | 0.67 | 0.74 | 0.74 | 0.74 | 0.73 | 0.71 | 0.72 | 0.56 |
| LMI | AUC | 0.65 | 0.65 | 0.65 | 0.64 | 0.62 | 0.74 | 0.69 | 0.73 | 0.70 | 0.74 | 0.73 | 0.72 | 0.72 | 0.57 |
| NM | AUC | 0.71 | 0.69 | 0.69 | 0.68 | 0.67 | 0.74 | 0.72 | 0.75 | 0.74 | 0.75 | 0.70 | 0.69 | 0.69 | 0.55 |
| NMI | AUC | 0.71 | 0.70 | 0.70 | 0.69 | 0.68 | 0.74 | 0.72 | 0.75 | 0.73 | 0.75 | 0.71 | 0.70 | 0.70 | 0.55 |
| | | **Dichotomous Outcome and Categorical ERS/ WQS/Q-gcomp** | | | | | | | | | | | | | |
| LM | OR | 11.6 | 10.6 | 10.9 | 10.1 | 9.4 | 11.7 | 8.2 | 11.2 | 12.0 | 11.4 | 9.4 | 7.0 | 9.5 | 1.5 |
| LMI | OR | 2.9 | 2.8 | 2.8 | 2.7 | 2.4 | 7.5 | 8.4 | 6.8 | 5.4 | 7.6 | 6.0 | 6.0 | 5.7 | 1.8 |
| NM | OR | 5.9 | 5.3 | 5.1 | 5.3 | 4.4 | 8.6 | 7.6 | 9.4 | 8.1 | 9.5 | 5.8 | 5.5 | 5.7 | 1.5 |
| NMI | OR | 6.3 | 4.9 | 5.0 | 5.3 | 4.5 | 7.7 | 7.2 | 8.8 | 7.5 | 8.8 | 6.0 | 6.0 | 5.9 | 1.5 |

*LM* linear main effects, *LMI* linear main effects and interactions, *NM* nonlinear main effects, *NMI* nonlinear main effects and interactions
*Corr* correlation, *SSE* sum of squared error, *AUC* area under the receiver operating characteristic curve, *OR* odds ratio
*Enet-M* elastic net for main effects, *WQS-M** weighted quantile sum regression (WQS) for selected main effects by Enet-M, *WQS-M* WQS for main effects, *Q-gcomp-M** quantile g-computation (Q-gcomp) for selected main effects by Enet-M, *Q-gcomp-M* Q-gcomp for main effects, *Enet-MI* elastic net for main effects and interactions, *BKMR* Bayesian kernel machine regression, *HierNet* lasso for hierarchical interactions, *SNIF* selection of nonlinear interactions by a forward stepwise algorithm, *SL* super learner, *WQS-MI** WQS for selected main effects and interactions by Enet-MI, *WQS-MI* WQS for main effects and interactions, *Q-gcomp-Mi** Q-gcomp for selected main effects and interactions by Enet-MI, *Q-gcomp-MI* Q-gcomp for main effects and interactions

**Table S7: Selection accuracy for main and interaction identification among five methods, where outcome is generated from Logit, LogitI, NLogitI, and NLogitI. Means of Sen, Spe, FDR and FPR are obtained from 100 data replications with $N_{train} = 1000$, $p = 40$, $q = 5$ and $R^2 = 0.2$**

| Data | Type | Metric | Lasso-M | Enet-M | G-Lasso-M | Lasso-MI | Enet-MI | G-Lasso-MI | RF | HierNet |
|---|---|---|---|---|---|---|---|---|---|---|
| Logit | Main/Marginal | Sen | 0.994 | 0.996 | 1.000 | 0.982 | 0.992 | 1.000 | 0.642 | 0.986 |
| | | Spe | 0.719 | 0.647 | 0.206 | 0.861 | 0.767 | 0.000 | 0.819 | 0.673 |
| | | FDR | 0.640 | 0.699 | 0.828 | 0.469 | 0.606 | 0.875 | 0.595 | 0.637 |
| | Interaction | FPR | -- | -- | -- | 0.021 | 0.032 | 1.000 | -- | 0.028 |
| LogitI | Main/Marginal | Sen | 0.968 | 0.974 | 1.000 | 0.914 | 0.952 | 1.000 | 0.598 | 0.968 |
| | | Spe | 0.752 | 0.674 | 0.077 | 0.872 | 0.803 | 0.000 | 0.747 | 0.585 |
| | | FDR | 0.613 | 0.682 | 0.857 | 0.457 | 0.568 | 0.875 | 0.706 | 0.688 |
| | Interaction | Sen | -- | -- | -- | 0.279 | 0.348 | 1.000 | -- | 0.316 |
| | | Spe | -- | -- | -- | 0.973 | 0.963 | 0.000 | -- | 0.971 |
| | | FDR | -- | -- | -- | 0.860 | 0.882 | 0.987 | -- | 0.839 |
| Nlogit | Main/Marginal | Sen | 0.612 | 0.662 | 0.990 | 0.542 | 0.576 | 0.990 | 0.364 | 0.866 |
| | | Spe | 0.821 | 0.730 | 0.169 | 0.923 | 0.858 | 0.010 | 0.861 | 0.627 |
| | | FDR | 0.627 | 0.724 | 0.839 | 0.443 | 0.606 | 0.866 | 0.570 | 0.667 |
| | Interaction | FPR | -- | -- | -- | 0.023 | 0.036 | 0.990 | -- | 0.026 |
| NlogitI | Main/Marginal | Sen | 0.722 | 0.764 | 0.990 | 0.584 | 0.622 | 1.000 | 0.374 | 0.946 |
| | | Spe | 0.792 | 0.704 | 0.186 | 0.913 | 0.833 | 0.000 | 0.849 | 0.618 |
| | | FDR | 0.641 | 0.718 | 0.831 | 0.467 | 0.631 | 0.875 | 0.611 | 0.685 |
| | Interaction | Sen | -- | -- | -- | 0.153 | 0.208 | 1.000 | -- | 0.140 |
| | | Spe | -- | -- | -- | 0.974 | 0.960 | 0.000 | -- | 0.974 |
| | | FDR | -- | -- | -- | 0.922 | 0.932 | 0.987 | -- | 0.924 |

*Logit* logit-link linear main effects, *LogitI* logit-link linear main effects and interactions, *Nlogit* logit-link nonlinear main effects, *NlogitI* logit-link nonlinear main effects and interactions
*Sen* sensitivity, *Spe* specificity, *FDR* false discovery rate, *FPR* false positive rate
*Lasso-M* lasso for main effects, *Enet-M* elastic net for main effects, *G-Lasso-M* group lasso for main effects, *Lasso-MI* lasso for main effects and interactions, *Enet-MI* elastic net for main effects and interactions, *G-Lasso-MI* group lasso for main effects and interactions, *RF* random forest, *HierNet* lasso for hierarchical interactions
Mean prevalence of outcome over 100 replicates equals to 13.8%, 12.8%, 14.5% and 11.2% for Logit, LogitI, Nlogit and NlogitI, respectively

**Table S8: Risk prediction performance by different statistical methods, when data are generated from Logit, LogitI, Nlogit, and NLogitI. Means of AUC, Brier, median of OR are obtained from 100 data replications for $N_{test} = 1000, p = 40, q = 5$ and $R^2 = 0.2$**

| Data | Metric | ERS Enet-M | WQS -M* | WQS -M | Q-gcomp -M* | Q-gcomp -M | ERS Lasso-MI | ERS Enet-MI | ERS RF | ERS HierNet | ERS SL | WQS -MI* | WQS -MI | Q-gcomp -MI* | Q-gcomp -MI |
|---|---|---|---|---|---|---|---|---|---|---|---|---|---|---|---|
| | | **Dichotomous Outcome and Continuous ERS WQS/Q-gcomp** | | | | | | | | | | | | | |
| Logit | AUC | 0.816 | 0.803 | 0.801 | 0.798 | 0.786 | 0.809 | 0.808 | 0.779 | 0.808 | 0.810 | 0.777 | 0.746 | 0.768 | 0.560 |
| | Brier | 0.097 | 0.100 | 0.100 | 0.101 | 0.104 | 0.098 | 0.099 | 0.102 | 0.099 | 0.099 | 0.103 | 0.104 | 0.108 | 0.361 |
| LogitI | AUC | 0.775 | 0.765 | 0.764 | 0.761 | 0.746 | 0.764 | 0.765 | 0.738 | 0.767 | 0.765 | 0.737 | 0.723 | 0.726 | 0.559 |
| | Brier | 0.094 | 0.096 | 0.097 | 0.097 | 0.100 | 0.094 | 0.094 | 0.097 | 0.094 | 0.094 | 0.097 | 0.099 | 0.103 | 0.357 |
| Nlogit | AUC | 0.756 | 0.741 | 0.736 | 0.731 | 0.714 | 0.765 | 0.762 | 0.739 | 0.761 | 0.765 | 0.731 | 0.711 | 0.715 | 0.539 |
| | Brier | 0.108 | 0.112 | 0.112 | 0.114 | 0.118 | 0.107 | 0.108 | 0.110 | 0.108 | 0.107 | 0.113 | 0.115 | 0.119 | 0.382 |
| NlogitI | AUC | 0.785 | 0.768 | 0.765 | 0.761 | 0.744 | 0.796 | 0.792 | 0.768 | 0.796 | 0.796 | 0.757 | 0.747 | 0.741 | 0.553 |
| | Brier | 0.085 | 0.088 | 0.088 | 0.090 | 0.093 | 0.084 | 0.084 | 0.087 | 0.083 | 0.084 | 0.088 | 0.089 | 0.095 | 0.351 |
| | | **Dichotomous Outcome and Categorical ERS WQS/Q-gcomp** | | | | | | | | | | | | | |
| Logit | OR | 33.1 | 31.4 | 28.0 | 27.4 | 25.9 | 24.0 | 28.1 | 27.8 | 17.7 | 28.3 | 30.0 | 14.9 | 8.8 | 16.4 |
| LogitI | OR | 10.7 | 10.7 | 10.3 | 10.2 | 9.0 | 9.4 | 9.5 | 4.8 | 9.7 | 9.2 | 7.3 | 6.4 | 6.7 | 1.5 |
| Nlogit | OR | 11.6 | 10.1 | 9.3 | 10.2 | 8.2 | 12.3 | 11.9 | 7.8 | 12.3 | 13.3 | 9.0 | 6.7 | 7.3 | 1.3 |
| NlogitI | OR | 15.5 | 12.2 | 12.2 | 12.8 | 11.3 | 17.0 | 18.0 | 9.1 | 16.4 | 18.3 | 12.1 | 10.3 | 9.4 | 1.5 |

*Logit* logit-link linear main effects, *LogitI* logit-link linear main effects and interactions, *Nlogit* logit-link nonlinear main effects, *NlogitI* logit-link nonlinear main effects and interactions
*AUC* area under the receiver operating characteristic curve, *Brier* Brier score, *OR* odds ratio
*Enet-M* elastic net for main effects, *WQS-M** weighted quantile sum regression (WQS) for selected main effects by Enet-M, *WQS-M* WQS for main effects, *Q-gcomp-M** quantile g-computation (Q-gcomp) for selected main effects by Enet-M, *Q-gcomp-M* Q-gcomp for main effects, *Lasso-MI* lasso for main effects and interactions, *Enet-MI* elastic net for main effects and interactions, *RF* random forest, *HierNet* lasso for hierarchical interactions, *SL* super learner, *WQS-MI** WQS for selected main effects and interactions by Enet-MI, *WQS-MI* WQS for main effects and interactions, *Q-gcomp-Mi** Q-gcomp for selected main effects and interactions by Enet-MI, *Q-gcomp-MI* Q-gcomp for main effects and interactions
Mean prevalence of outcome over 100 replicates equals to 13.8%, 12.8%, 14.5% and 11.2% for Logit, LogitI, Nlogit and NlogitI, respectively

**Table S9: Selection accuracy for main and interaction identification among eight methods, where outcome is generated from LM, NM, LMI, and NMI. Means of Sen, Spe, FDR and FPR are obtained from 100 data replications with $N_{train} = 1000$, $p = 40$, $q = 5$, and $R^2 = 0.1$**

| Data | Type | Metric | Lasso-M | Enet-M | Lasso-MI | Enet-MI | BKMR | RF | HierNet | SNIF |
|---|---|---|---|---|---|---|---|---|---|---|
| LM | Main/Marginal | Sen | 0.926 | 0.926 | 0.882 | 0.898 | 0.992 | 0.614 | 0.922 | 0.622 |
| | | Spe | 0.729 | 0.719 | 0.843 | 0.820 | 0.004 | 0.865 | 0.697 | 0.973 |
| | | FDR | 0.641 | 0.654 | 0.535 | 0.565 | 0.875 | 0.517 | 0.671 | 0.211 |
| | Interaction | FPR | -- | -- | 0.014 | 0.016 | -- | -- | 0.022 | 0.000 |
| LMI | Main/Marginal | Sen | 0.552 | 0.582 | 0.502 | 0.506 | 0.996 | 0.632 | 0.950 | 0.410 |
| | | Spe | 0.788 | 0.775 | 0.869 | 0.858 | 0.006 | 0.785 | 0.397 | 0.962 |
| | | FDR | 0.695 | 0.706 | 0.632 | 0.648 | 0.875 | 0.665 | 0.808 | 0.334 |
| | Interaction | Sen | -- | -- | 0.469 | 0.488 | -- | -- | 0.361 | 0.045 |
| | | Spe | -- | -- | 0.967 | 0.964 | -- | -- | 0.966 | 1.000 |
| | | FDR | -- | -- | 0.833 | 0.843 | -- | -- | 0.868 | 0.145 |
| NM | Main/Marginal | Sen | 0.548 | 0.546 | 0.472 | 0.500 | 0.996 | 0.482 | 0.852 | 0.590 |
| | | Spe | 0.783 | 0.784 | 0.900 | 0.886 | 0.019 | 0.916 | 0.661 | 0.981 |
| | | FDR | 0.667 | 0.685 | 0.550 | 0.579 | 0.873 | 0.343 | 0.716 | 0.135 |
| | Interaction | FPR | -- | -- | 0.028 | 0.032 | -- | -- | 0.022 | 0.000 |
| NMI | Main/Marginal | Sen | 0.586 | 0.584 | 0.500 | 0.520 | 1.000 | 0.536 | 0.888 | 0.590 |
| | | Spe | 0.784 | 0.791 | 0.888 | 0.872 | 0.009 | 0.871 | 0.621 | 0.979 |
| | | FDR | 0.672 | 0.671 | 0.570 | 0.604 | 0.874 | 0.449 | 0.734 | 0.160 |
| | Interaction | Sen | -- | -- | 0.171 | 0.188 | -- | -- | 0.129 | 0.007 |
| | | Spe | -- | -- | 0.971 | 0.966 | -- | -- | 0.976 | 1.000 |
| | | FDR | -- | -- | 0.926 | 0.930 | -- | -- | 0.931 | 0.020 |

*LM* linear main effects, *LMI* linear main effects and interactions, *NM* nonlinear main effects, *NMI* nonlinear main effects and interactions
*Sen* sensitivity, *Spe* specificity, *FDR* false discovery rate, *FPR* false positive rate
*Lasso-M* lasso for main effects, *Enet-M* elastic net for main effects, *Lasso-MI* lasso for main effects and interactions, *Enet-MI* elastic net for main effects and interactions, *BKMR* Bayesian kernel machine regression, *RF* random forest, *HierNet* lasso for hierarchical interactions, *SNIF* selection of nonlinear interactions by a forward stepwise algorithm

**Table S10: Risk prediction performance by different statistical methods, when data are generated from LM, NM, LMI, and NMI. Means of Corr, SSE, AUC, and median of OR are obtained from 100 data replications for $N_{test} = 1000$, $p = 40$, $q = 5$, and $R^2 = 0.1$**

| Data | Metric | ERS Enet-M | WQS -M* | WQS -M | Q-gcomp -M* | Q-gcomp -M | ERS Enet-MI | ERS BKMR | ERS HierNet | ERS SNIF | ERS SL | WQS -MI* | WQS -MI | Q-gcomp -MI* | Q-gcomp -MI |
|---|---|---|---|---|---|---|---|---|---|---|---|---|---|---|---|
| | | **Continuous Outcome and Continuous ERS/WQS/Q-gcomp** | | | | | | | | | | | | | |
| LM | Corr | 0.29 | 0.28 | 0.28 | 0.27 | 0.25 | 0.28 | 0.15 | 0.28 | 0.27 | 0.28 | 0.26 | 0.21 | 0.25 | 0.04 |
| | SSE | 81.9 | 82.6 | 82.5 | 83.2 | 84.8 | 82.5 | 88.6 | 82.4 | 82.9 | 82.3 | 83.8 | 85.7 | 85.5 | 540.8 |
| LMI | Corr | 0.12 | 0.12 | 0.13 | 0.11 | 0.09 | 0.26 | 0.10 | 0.24 | 0.17 | 0.26 | 0.23 | 0.23 | 0.21 | 0.04 |
| | SSE | 356.9 | 358.0 | 357.0 | 360.8 | 369.0 | 337.7 | 366.3 | 342.2 | 358.6 | 338.2 | 345.5 | 343.5 | 359.3 | 2268.5 |
| NM | Corr | 0.21 | 0.20 | 0.19 | 0.18 | 0.15 | 0.25 | 0.12 | 0.28 | 0.26 | 0.28 | 0.21 | 0.19 | 0.19 | 0.03 |
| | SSE | 154.7 | 155.9 | 156.6 | 157.3 | 161.1 | 152.0 | 170.4 | 149.7 | 157.7 | 149.6 | 155.8 | 156.4 | 161.2 | 1013.4 |
| NMI | Corr | 0.20 | 0.19 | 0.19 | 0.18 | 0.15 | 0.25 | 0.11 | 0.27 | 0.24 | 0.27 | 0.21 | 0.21 | 0.19 | 0.03 |
| | SSE | 233.3 | 235.0 | 235.3 | 236.5 | 242.5 | 228.2 | 255.6 | 226.2 | 236.0 | 226.1 | 233.7 | 233.2 | 242.0 | 1521.3 |
| | | **Dichotomous Outcome and Continuous ERS/ WQS/Q-gcomp** | | | | | | | | | | | | | |
| LM | AUC | 0.66 | 0.66 | 0.66 | 0.65 | 0.64 | 0.66 | 0.59 | 0.66 | 0.65 | 0.66 | 0.65 | 0.62 | 0.64 | 0.53 |
| LMI | AUC | 0.59 | 0.59 | 0.60 | 0.59 | 0.57 | 0.65 | 0.58 | 0.64 | 0.61 | 0.65 | 0.64 | 0.64 | 0.63 | 0.53 |
| NM | AUC | 0.64 | 0.63 | 0.63 | 0.62 | 0.60 | 0.65 | 0.58 | 0.66 | 0.65 | 0.66 | 0.63 | 0.62 | 0.62 | 0.53 |
| NMI | AUC | 0.64 | 0.63 | 0.63 | 0.62 | 0.60 | 0.65 | 0.57 | 0.66 | 0.65 | 0.66 | 0.63 | 0.63 | 0.62 | 0.53 |
| | | **Dichotomous Outcome and Categorical ERS/ WQS/Q-gcomp** | | | | | | | | | | | | | |
| LM | OR | 4.8 | 4.4 | 4.5 | 4.1 | 3.6 | 4.7 | 3.1 | 4.6 | 4.3 | 4.8 | 4.0 | 3.1 | 3.7 | 1.2 |
| LMI | OR | 2.0 | 2.0 | 2.1 | 1.9 | 1.7 | 3.5 | 2.9 | 3.1 | 2.5 | 3.7 | 3.3 | 3.0 | 2.9 | 1.3 |
| NM | OR | 3.2 | 3.1 | 3.0 | 2.9 | 2.4 | 3.7 | 2.7 | 4.0 | 3.6 | 4.0 | 3.0 | 2.8 | 3.0 | 1.2 |
| NMI | OR | 3.1 | 3.1 | 2.8 | 2.7 | 2.4 | 3.7 | 2.8 | 3.7 | 3.4 | 3.9 | 3.0 | 3.0 | 2.9 | 1.2 |

*LM* linear main effects, *LMI* linear main effects and interactions, *NM* nonlinear main effects, *NMI* nonlinear main effects and interactions
*Corr* correlation, *SSE* sum of squared error, *AUC* area under the receiver operating characteristic curve, *OR* odds ratio
*Enet-M* elastic net for main effects, *WQS-M** weighted quantile sum regression (WQS) for selected main effects by Enet-M, *WQS-M* WQS for main effects, *Q-gcomp-M** quantile g-computation (Q-gcomp) for selected main effects by Enet-M, *Q-gcomp-M* Q-gcomp for main effects, *Enet-MI* elastic net for main effects and interactions, *BKMR* Bayesian kernel machine regression, *HierNet* lasso for hierarchical interactions, *SNIF* selection of nonlinear interactions by a forward stepwise algorithm, *SL* super learner, *WQS-MI** WQS for selected main effects and interactions by Enet-MI, *WQS-MI* WQS for main effects and interactions, *Q-gcomp-Mi** Q-gcomp for selected main effects and interactions by Enet-MI, *Q-gcomp-MI* Q-gcomp for main effects and interactions

**Table S11: Selection accuracy for main and interaction identification among five methods, where outcome is generated from Logit, LogitI, NLogitI, and NLogitI. Means of Sen, Spe, FDR and FPR are obtained from 100 data replications with $N_{train} = 1000, p = 40, q = 5$ and $R^2 = 0.1$**

| Data | Type | Metric | Lasso-M | Enet-M | G-Lasso-M | Lasso-MI | Enet-MI | G-Lasso-MI | RF | HierNet |
|---|---|---|---|---|---|---|---|---|---|---|
| Logit | Main/Marginal | Sen | 0.920 | 0.940 | 0.910 | 0.854 | 0.898 | 1.000 | 0.608 | 0.740 |
| | | Spe | 0.774 | 0.699 | 0.236 | 0.863 | 0.806 | 0.000 | 0.745 | 0.773 |
| | | FDR | 0.609 | 0.672 | 0.787 | 0.499 | 0.582 | 0.875 | 0.693 | 0.521 |
| | Interaction | FPR | -- | -- | -- | 0.016 | 0.023 | 1.000 | -- | 0.018 |
| LogitI | Main/Marginal | Sen | 0.838 | 0.866 | 0.960 | 0.764 | 0.810 | 0.980 | 0.490 | 0.772 |
| | | Spe | 0.786 | 0.720 | 0.139 | 0.880 | 0.831 | 0.020 | 0.739 | 0.700 |
| | | FDR | 0.619 | 0.674 | 0.809 | 0.495 | 0.573 | 0.858 | 0.762 | 0.614 |
| | Interaction | Sen | -- | -- | -- | 0.112 | 0.147 | 0.980 | -- | 0.137 |
| | | Spe | -- | -- | -- | 0.981 | 0.974 | 0.020 | -- | 0.979 |
| | | FDR | -- | -- | -- | 0.918 | 0.926 | 0.967 | -- | 0.759 |
| Nlogit | Main/Marginal | Sen | 0.598 | 0.642 | 0.960 | 0.524 | 0.550 | 1.000 | 0.378 | 0.768 |
| | | Spe | 0.824 | 0.735 | 0.207 | 0.929 | 0.868 | 0.000 | 0.829 | 0.687 |
| | | FDR | 0.632 | 0.717 | 0.809 | 0.429 | 0.600 | 0.875 | 0.652 | 0.653 |
| | Interaction | FPR | -- | -- | -- | 0.020 | 0.029 | 1.000 | -- | 0.021 |
| NlogitI | Main/Marginal | Sen | 0.606 | 0.656 | 0.960 | 0.520 | 0.560 | 0.990 | 0.370 | 0.778 |
| | | Spe | 0.829 | 0.755 | 0.181 | 0.921 | 0.870 | 0.010 | 0.826 | 0.661 |
| | | FDR | 0.622 | 0.702 | 0.823 | 0.462 | 0.590 | 0.866 | 0.663 | 0.635 |
| | Interaction | Sen | -- | -- | -- | 0.072 | 0.100 | 0.990 | -- | 0.095 |
| | | Spe | -- | -- | -- | 0.980 | 0.969 | 0.010 | -- | 0.978 |
| | | FDR | -- | -- | -- | 0.946 | 0.954 | 0.977 | -- | 0.843 |

*Logit* logit-link linear main effects, *LogitI* logit-link linear main effects and interactions, *Nlogit* logit-link nonlinear main effects, *NlogitI* logit-link nonlinear main effects and interactions

*Sen* sensitivity, *Spe* specificity, *FDR* false discovery rate, *FPR* false positive rate

*Lasso-M* lasso for main effects, *Enet-M* elastic net for main effects, *G-Lasso-M* group lasso for main effects, *Lasso-MI* lasso for main effects and interactions, *Enet-MI* elastic net for main effects and interactions, *G-Lasso-MI* group lasso for main effects and interactions, *RF* random forest, *HierNet* lasso for hierarchical interactions

Mean prevalence of outcome over 100 replicates equals to 15.3%, 15.1%, 16.4% and 15.4% for Logit, LogitI, Nlogit and NlogitI, respectively

**Table S12: Risk prediction performance by different statistical methods, when data are generated from Logit, LogitI, Nlogit, and NLogitI. Means of AUC, Brier, and median of OR are obtained from 100 data replications for $N_{test} = 1000, p = 40, q = 5$ and $R^2 = 0.1$**

| Data | Metric | ERS Enet-M | WQS -M* | WQS -M | Q-gcomp -M* | Q-gcomp -M | ERS Lasso-MI | ERS Enet-MI | ERS RF | ERS HierNet | ERS SL | WQS -MI* | WQS -MI | Q-gcomp -MI* | Q-gcomp -MI |
|---|---|---|---|---|---|---|---|---|---|---|---|---|---|---|---|
| | | **Dichotomous Outcome and Continuous ERS WQS/Q-gcomp** | | | | | | | | | | | | | |
| Logit | AUC | 0.720 | 0.712 | 0.711 | 0.706 | 0.689 | 0.711 | 0.712 | 0.682 | 0.697 | 0.712 | 0.683 | 0.653 | 0.680 | 0.531 |
| | Brier | 0.120 | 0.121 | 0.121 | 0.122 | 0.126 | 0.121 | 0.121 | 0.123 | 0.123 | 0.122 | 0.123 | 0.125 | 0.127 | 0.391 |
| LogitI | AUC | 0.685 | 0.677 | 0.681 | 0.674 | 0.655 | 0.671 | 0.672 | 0.647 | 0.670 | 0.673 | 0.651 | 0.635 | 0.643 | 0.528 |
| | Brier | 0.121 | 0.122 | 0.122 | 0.123 | 0.126 | 0.121 | 0.121 | 0.124 | 0.122 | 0.121 | 0.123 | 0.123 | 0.128 | 0.391 |
| Nlogit | AUC | 0.714 | 0.701 | 0.697 | 0.692 | 0.674 | 0.716 | 0.714 | 0.690 | 0.714 | 0.716 | 0.685 | 0.665 | 0.674 | 0.528 |
| | Brier | 0.126 | 0.128 | 0.128 | 0.130 | 0.134 | 0.126 | 0.126 | 0.128 | 0.126 | 0.126 | 0.130 | 0.131 | 0.135 | 0.400 |
| NlogitI | AUC | 0.709 | 0.695 | 0.691 | 0.690 | 0.670 | 0.710 | 0.708 | 0.682 | 0.705 | 0.710 | 0.680 | 0.664 | 0.668 | 0.529 |
| | Brier | 0.120 | 0.122 | 0.122 | 0.123 | 0.127 | 0.120 | 0.120 | 0.123 | 0.121 | 0.120 | 0.123 | 0.124 | 0.128 | 0.393 |
| | | **Dichotomous Outcome and Categorical ERS WQS/Q-gcomp** | | | | | | | | | | | | | |
| Logit | OR | 8.7 | 7.7 | 7.8 | 7.4 | 6.4 | 7.9 | 7.9 | 5.0 | 6.9 | 8.1 | 5.4 | 3.7 | 5.5 | 1.3 |
| LogitI | OR | 5.2 | 4.9 | 4.9 | 4.8 | 4.2 | 4.5 | 4.4 | 2.9 | 4.3 | 4.7 | 3.7 | 3.2 | 3.5 | 1.2 |
| Nlogit | OR | 7.4 | 6.4 | 6.1 | 6.8 | 5.3 | 7.7 | 7.8 | 5.1 | 7.9 | 8.3 | 5.5 | 4.6 | 4.9 | 1.2 |
| NlogitI | OR | 7.0 | 5.9 | 5.7 | 6.0 | 4.8 | 7.2 | 6.9 | 4.5 | 6.8 | 7.8 | 5.2 | 4.2 | 4.3 | 1.2 |

*Logit* logit-link linear main effects, *LogitI* logit-link linear main effects and interactions, *Nlogit* logit-link nonlinear main effects, *NlogitI* logit-link nonlinear main effects and interactions
*AUC* area under the receiver operating characteristic curve, *Brier* Brier score, *OR* odds ratio
*Enet-M* elastic net for main effects, *WQS-M** weighted quantile sum regression (WQS) for selected main effects by Enet-M, *WQS-M* WQS for main effects, *Q-gcomp-M** quantile g-computation (Q-gcomp) for selected main effects by Enet-M, *Q-gcomp-M* Q-gcomp for main effects, *Lasso-MI* lasso for main effects and interactions, *Enet-MI* elastic net for main effects and interactions, *RF* random forest, *HierNet* lasso for hierarchical interactions, *SL* super learner, *WQS-MI** WQS for selected main effects and interactions by Enet-MI, *WQS-MI* WQS for main effects and interactions, *Q-gcomp-Mi** Q-gcomp for selected main effects and interactions by Enet-MI, *Q-gcomp-MI* Q-gcomp for main effects and interactions
Mean prevalence of outcome over 100 replicates equals to 15.3%, 15.1%, 16.4% and 15.4% for Logit, LogitI, Nlogit and NlogitI, respectively

**Table S13: Mean computing time (seconds) over 100 data replications to fit the training data under settings of $N_{train} = 500$ and $p = 20$; $N_{train} = 1,000$ and $p = 40$ for different methods, $R^2 = 0.1$ for all settings of continuous and binary outcomes**

| $N_{train}$ | Data | WQS-M | Q-gcomp-M | Lasso-MI | Enet-MI | G-Lasso-MI | BKMR | RF | HigLasso | HierNet | SNIF |
|---|---|---|---|---|---|---|---|---|---|---|---|
| 500 | LM | 6.5 | 0.01 | 0.7 | 0.6 | 345.0 | 2056.1 | 1.3 | 9001.0 | 43.0 | 1.8 |
| | LMI | 6.8 | 0.01 | 0.7 | 0.6 | 389.9 | 2061.8 | 1.3 | 52233.9 | 62.6 | 1.8 |
| | NM | 6.6 | 0.01 | 0.7 | 0.6 | 370.6 | 1649.5 | 1.3 | 28355.0 | 50.2 | 2.4 |
| | NMI | 6.8 | 0.01 | 0.7 | 0.6 | 385.7 | 1868.5 | 1.4 | 47837.8 | 53.8 | 2.3 |
| 1,000 | LM | 23.5 | 0.03 | 27.4 | 26.6 | -- | 33195.0 | 6.0 | -- | 352.6 | 14.3 |
| | LMI | 24.9 | 0.03 | 26.2 | 26.2 | -- | 33032.3 | 6.6 | -- | 528.0 | 17.2 |
| | NM | 24.2 | 0.03 | 23.5 | 23.0 | -- | 31201.0 | 6.3 | -- | 394.5 | 21.5 |
| | NMI | 25.2 | 0.03 | 21.1 | 20.7 | -- | 31038.7 | 6.3 | -- | 410.5 | 20.5 |
| 500 | Logit | 8.0 | 0.02 | 9.6 | 8.7 | 189.4 | -- | 0.8 | -- | 44.4 | -- |
| | LogitI | 7.9 | 0.01 | 10.7 | 9.6 | 171.1 | -- | 0.9 | -- | 49.5 | -- |
| | Nlogit | 7.8 | 0.01 | 10.9 | 9.6 | 206.5 | -- | 0.9 | -- | 39.7 | -- |
| | NlogitI | 7.9 | 0.01 | 9.9 | 8.9 | 183.2 | -- | 0.9 | -- | 42.4 | -- |
| 1,000 | Logit | 34.0 | 0.03 | 22.1 | 22.0 | 2888.3 | -- | 3.5 | -- | 328.6 | -- |
| | LogitI | 32.6 | 0.04 | 22.7 | 22.8 | 1655.9 | -- | 3.6 | -- | 358.6 | -- |
| | Nlogit | 32.4 | 0.04 | 23.4 | 23.0 | 3027.8 | -- | 3.6 | -- | 286.7 | -- |
| | NlogitI | 32.0 | 0.03 | 23.4 | 23.1 | 2566.2 | -- | 3.8 | -- | 323.7 | -- |

*WQS-M* WQS for main effects, *Q-gcomp-M* Q-gcomp for main effects, *Lasso-MI* lasso for main effects and interactions, *Enet-MI* elastic net for main effects and interactions, *G-Lasso-MI* group lasso for main effects and interactions, *BKMR* Bayesian kernel machine regression, *RF* random forest, *HigLasso* hierarchical integrative group lasso, *HierNet* lasso for hierarchical interactions, *SNIF* selection of nonlinear interactions by a forward stepwise algorithm
